# Development and assessment of a machine learning tool for predicting emergency admission in Scotland

**DOI:** 10.1101/2021.08.06.21261593

**Authors:** James Liley, Gergo Bohner, Samuel R. Emerson, Bilal A. Mateen, Katie Borland, David Carr, Scott Heald, Samuel D. Oduro, Jill Ireland, Keith Moffat, Rachel Porteous, Stephen Riddell, Simon Rogers, Ioanna Thoma, Nathan Cunningham, Chris Holmes, Katrina Payne, Sebastian J. Vollmer, Catalina A. Vallejos, Louis J. M. Aslett

**Affiliations:** Alan Turing Institute, London, UK; MRC Human Genetics Unit, Institute of Genetics and Cancer, University of Edinburgh, UK; Department of Mathematical Sciences, Durham University, UK; Mathematics Institute, University of Warwick, UK; Institute of Health Informatics, University College London, UK, and Wellcome Trust, London, UK; Public Health Scotland (PHS); former employee; University of St Andrews, UK; NHS National Services Scotland, UK; Department of Statistics, University of Warwick, UK; Department of Statistics, University of Oxford, UK

**Keywords:** Emergency admission, Primary care, Machine learning

## Abstract

Emergency admissions (EA), where a patient requires urgent in-hospital care, are a major challenge for healthcare systems. The development of risk prediction models can partly alleviate this problem by supporting primary care interventions and public health planning. Here, we introduce SPARRA*v*4, a predictive score for EA risk that will be deployed nationwide in Scotland. SPARRA*v*4 was derived using supervised and unsupervised machine-learning methods applied to routinely collected electronic health records from approximately 4.8M Scottish residents (2013-18). We demonstrate improvements in discrimination and calibration with respect to previous scores deployed in Scotland, as well as stability over a 3-year timeframe. Our analysis also provides insights about the epidemiology of EA risk in Scotland, by studying predictive performance across different population sub-groups and reasons for admission, as well as by quantifying the effect of individual input features. Finally, we discuss broader challenges including reproducibility and how to safely update risk prediction models that are already deployed at population level.

## Introduction

Emergency admissions (EA), where a patient requires urgent in-hospital care, represent deteriorations in individual health and are a major challenge for healthcare systems. For example, approximately 395,000 Scottish residents (≈ 1 in 14) had at least one EA between 1 April 2021 and 31 March 2022 [Public Health Scotland, 2022]. In total, around 600,000 EAs were recorded for these individuals, nearly 54% of all hospital admissions in that period, and they resulted in longer hospital stays (6.8 days average) compared to planned elective admissions (3.6 days average). Modern health and social care policies aim to implement proactive strategies [Rural Access Action Team, 2005], often by appropriate primary care intervention [McDonagh et al., 2000, Sanderson and Dixon, 2000, Coast et al., 1996]. Machine learning (ML) can support such interventions by identifying individuals at risk of EA who may benefit from anticipatory care. If successful, such interventions can be expected to improve patient outcomes and reduced pressures on secondary care (Figure 1A).

**Figure 1:**
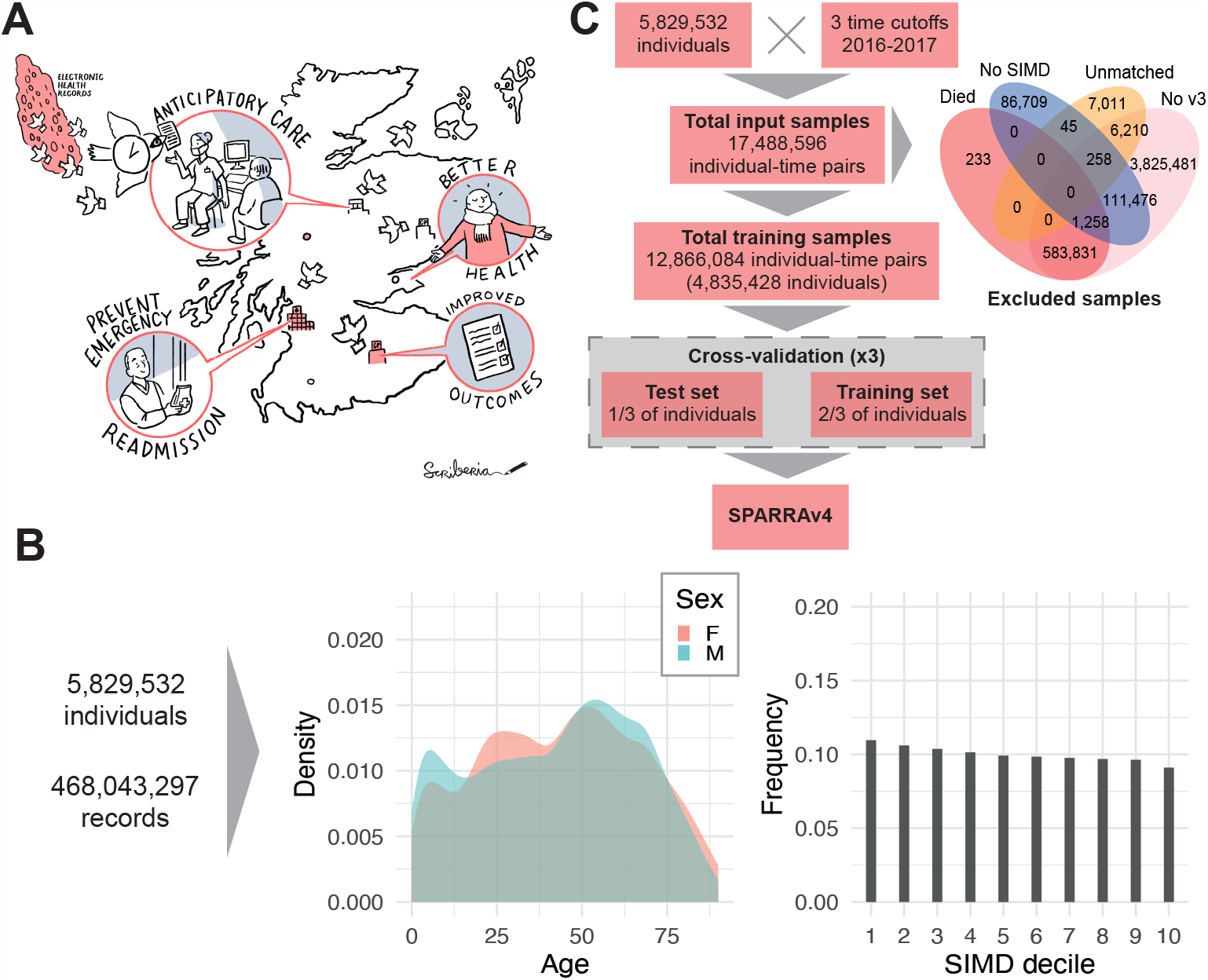
Data and model fitting overview. (A) Illustration of how SPARRA can support primary care intervention with the goal of improving patient outcomes. (B) Distribution of the number of input EHR entries (prior to exclusions) according to age, sex and SIMD deciles (1: most deprived; 10: least deprived). (C) Flow chart summarising data and model fitting pipelines.

A range of risk prediction models have been developed in this context [Rahimian et al., 2018, Lyon et al., 2007, Wallace et al., 2014, Bottle et al., 2006, Billings et al., 2006, Hippisley-Cox and Coupland, 2013]. However, transferability across temporal and geographical settings is limited due to differing demographics and data availability [Wallace et al., 2014]. Development of models in the setting in which they will be used is thus preferable to reapplication of models trained in other settings. In Scotland, the Information Services Division of the National Services Scotland (now incorporated into Public Health Scotland; PHS) developed SPARRA (Scottish Patients At Risk of Re-admission and Admission) — an algorithm to predict the risk of EA in the next 12 months. SPARRA was derived using national electronic health records (EHR) databases and has been in use since 2006. The current version of the algorithm (SPARRA*v*3) [Health and Social Care Information Programme, 2011] was deployed in 2012/13 and is calculated monthly by PHS for almost the entire Scottish population. Individual-level SPARRA scores can be accessed by general practitioners (GPs), helping them to plan mitigation strategies for individuals with complex care needs. Collectively, SPARRA scores may be used to estimate future demand, supporting planning and resource allocation. SPARRA has also been used extensively in public health research [Leckcivilize et al., 2021, Highet et al., 2014, Bajaj et al., 2016, Canny et al., 2016, Manoukian et al., 2021, Wallace et al., 2016].

In this paper we update the SPARRA algorithm to version 4 (SPARRA*v*4) using contemporary supervised and unsupervised ML methods. In particular, we use an ensemble of machine learning methods [Van der Laan et al., 2007], and use a topic model [Blei et al., 2003] to derive further information from prescriptions and diagnostic data. This represents a large scale ML risk score, fitted and deployed at national level, and widely available in clinical settings. We develop SPARRA*v*4 using EHRs collected for around 4.8 million (after exclusions) Scottish residents between 2013 and 2018. Among other variables, this includes data about past hospital admissions, long term conditions (e.g. asthma) and prescriptions. We use cross-validation to evaluate the validity of SPARRA*v*4 and its stability over time. This shows an improvement of performance with respect to SPARRA*v*3 in terms of discrimination and calibration, including a stratified analysis across different subpopulations. We also perform extensive analyses to determine what reasons for emergency admission are predictable, and use Shapley values [Lundberg and Lee, 2017] to quantify the effect of individual input factors. Finally, we discuss some of the practical challenges that arise when developing and deploying models of this kind, including issues associated to updating risk scores that are already deployed at population level.

Reproducibility is critical to ensure reliable application of ML in clinical settings [McDermott et al., 2021]. To provide a transparent description of our pipeline, this manuscript conforms to the TRIPOD guidelines [Collins et al., 2015] (S1). Moreover, all code is publicly available at github.com/jamesliley/SPARRA*v*4. This includes non-disclosive outputs used to generate all the figures and tables presented in this article.

## Results

### Data overview

The input data prior to any exclusions combines multiple national EHR databases held by Public Health Scotland for 5.8 million Scottish residents between 1 May 2013 and 30 April 2018 (Supplementary Table S2), some of whom died during the observation period. These comprised 468 million records, comprising interactions with the Scottish healthcare system and deaths. The number of total available records varies across sex, age, and SIMD (Figure 1B), and when records are grouped by database (Supplementary Figure S1A). In particular, marginally more records are available for individuals in the most deprived areas (as measured by deciles of the 2016 Scottish Index of Multiple Deprivation (SIMD); [Scottish Government, 2016]), particularly within accidents and emergency and mental health hospital records. Two additional tables (see Supplementary Table S2) containing historic data about long term conditions (LTC, back to 1981) and mortality records were also used as input.

We selected three time cutoffs for model fitting (1 May and 1 December 2016, and 1 May 2017) leading to 17.4 million individual-time pairs, hereafter referred to as samples (Figure 1C). This choice was informed by the extent of data required to define the input features used by the score (3 years prior the time cutoff) and the prediction target (1 year after the time cutoff). We used the earliest (1 May 2016) and latest (1 May 2017) possible time cutoffs, and a third time cutoff halfway between these. Although we could have used more than one time cutoff between the earliest and latest, we deemed that this would add little because, for most patients, we expect to have negiblible variation in their input features and EA status from month to month. After exclusions (which were predominantly due to samples without SPARRA v3 scores; see Methods), the data comprise 12.8 million samples corresponding to 4.8 million individuals. Overall, the study cohort is slightly older, has more females, and is moderately more deprived than the general population (Table 1). The prediction target was defined as a recorded EA to a Scottish hospital or death in the year following the time cutoff (see Methods). In total, 1,142,169 EA or death events (9%) were observed across all samples. This includes 57,183 samples for which a death was recorded (without a prior EA within that year) and 1,084,986 samples for which an EA was recorded (amongst those, 107,827 deaths were observed after the EA). As expected, the proportion of deaths amongst the observed events increases with age (Supplementary Figure S1B). Moreover, patients with an EA or death event (in at least one time cutoff) are, on average, older and more deprived than those without an event (Table 1).

**Table 1:** Demographic summary for the different cohorts: the whole Scottish population (approximately 5.8 million), those present in the input databases at least one (17,488,596 samples comprising 5,829,532 unique individuals), our study cohort after exclusions (12,866,084 samples comprising 4,835,428 unique individuals) and our study cohort after stratifying by event status (EA or death: 1,142,169 samples comprising 667,566 unique individuals; no EA or death: 11,723,915 samples comprising 4,670,756 unique individuals). Summary statistics were calculated using sample-level data. The EA or death cohort includes individual-time pairs for which the individual at least one EA or died during the year after the time. LTC denotes long-term conditions (e.g. epilepsy). Data for the Scottish population is from the 2011 Census [Office for National Statistics et al., 2011].

| Variable | Scottish<br>population | Input<br>data | Cohort |  |  |
| --- | --- | --- | --- | --- | --- |
|  |  |  | After<br>exclusions | EA or<br>death | No EA or<br>death |
| Sex (%) |  |  |  |  |  |
| Male | 48.5 | 48.2 | 45.4 | 46.2 | 45.3 |
| Female | 51.5 | 51.8 | 54.6 | 53.7 | 54.7 |
| Age at time cutoff (%) |  |  |  |  |  |
| 0-19 | 16.9 | 21.1 | 19.6 | 11.8 | 20.4 |
| 20-70 | 71.2 | 64.2 | 64.9 | 50.1 | 66.4 |
| 71+ | 11.9 | 14.7 | 15.4 | 38.1 | 13.2 |
| SIMD decile (%) |  |  |  |  |  |
| 1-5 | 50.0 | 50.8 | 52.0 | 59.5 | 51.2 |
| 6-10 | 50.0 | 49.2 | 48.0 | 40.5 | 48.8 |
| Any LTC (%) | Unknown | 29.4 | 32.1 | 58.8 | 29.5 |

### Overall predictive performance

In held out test data, SPARRA*v*4 was effective at predicting EA, and outperformed SPARRA*v*3 on the basis of area-under-receiver-operator-characteristic (AUROC) and area-under-precisionrecall-curve (AUPRC) (Figure 2A-B). SPARRA*v*4 was also better calibrated, particularly for samples with observed risk ≈ 0.5 (Figure 2C). Whilst SPARRA*v*3 and SPARRA*v*4 scores were highly correlated, large discrepancies were observed for some samples (Supplementary Figure S2). In samples for whom *v*3 and *v*4 disagreed (defined as |*v*3−*v*4| *>* 0.1), we found that *v*4 was better-calibrated than *v*3 (Figure 2D).

**Figure 2:**
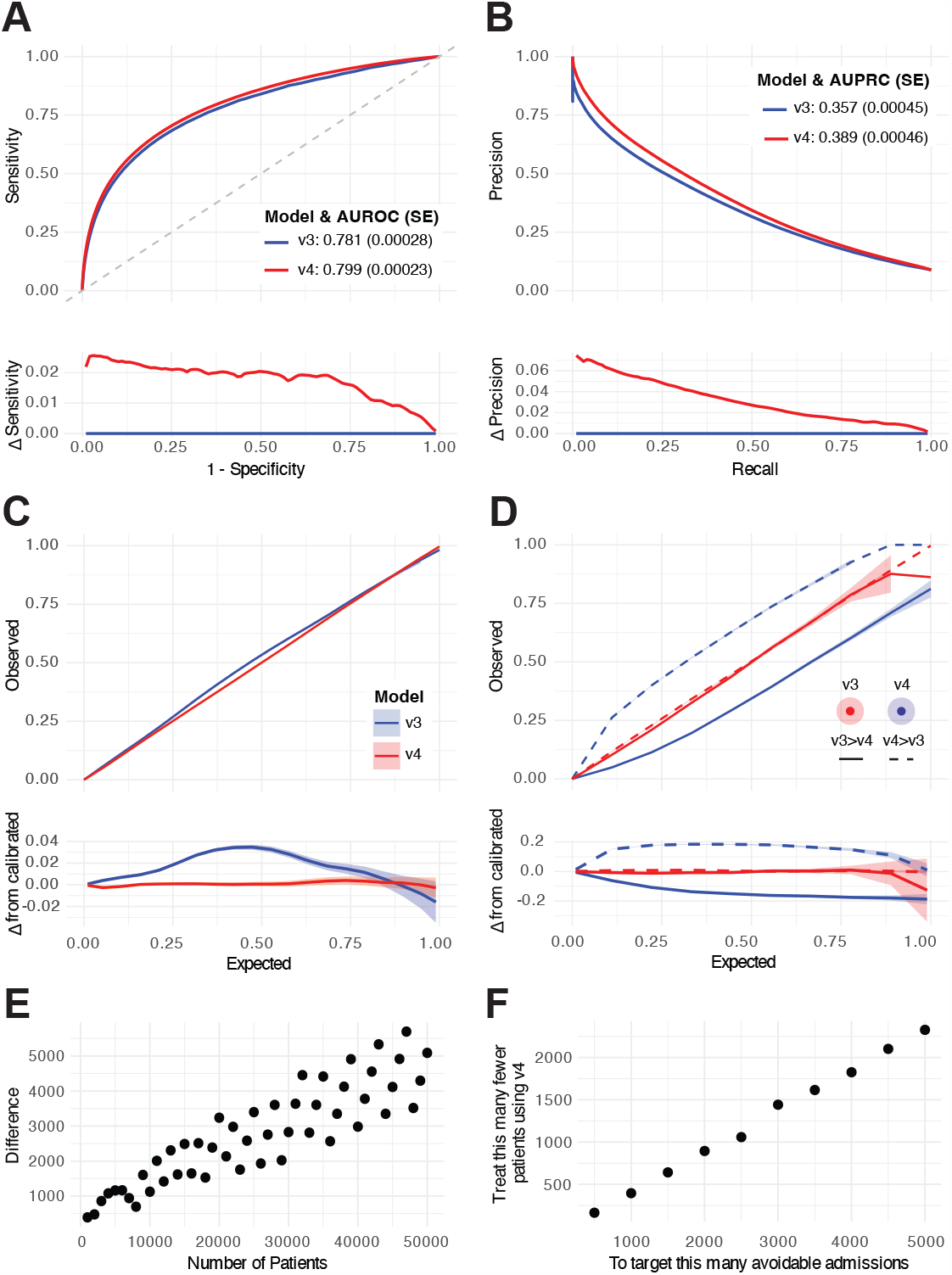
Comparison of overall predictive performance between SPARRA*v*3 and SPARRA*v*4. (A) ROC. (B) PRC. Lower sub-panels show differences in sensitivity and precision, respectively. Confidence intervals are negligible. (C) Calibration curves. (D) Calibration curves for samples in which |*v*4− *v*3 |*>* 0.1. Lower sub-panels show the difference between curves and the *y* = *x* line (perfect calibration). Confidence envelopes are pointwise (that is, for each *x*-value, not the whole curve). Predicted/true value pairs are combined across cross-validation folds in all panels for simplicity. (E) Difference in the number of individuals who had an event amongst individuals designated highest-risk by v3 and v4. The repeating pattern is a rounding effect of v3. (F) Difference in the number of highest-risk individuals to target to avoid a given number of admissions.

We also assessed the potential population-wide benefit of SPARRA*v*4 over SPARRA*v*3 directly. Amongst the 50,000 individuals judged to be at highest risk by SPARRA*v*3, around 4,000 fewer individuals were eventually admitted that were amongst the 50,000 individuals judged to be at highest risk by SPARRA*v*4 (Figure 2E). For another perspective, if we simply assume that 20% of admissions are avoidable [value taken from Blunt, 2013], that avoidable admissions are as predictable as non-avoidable admissions, and that we wish to pre-empt 3,000 avoidable admissions by targeted intervention on the highest risk patients (the second assumption is conservative, since avoidable admissions are often predictable due to other medical problems). Then, by using SPARRA*v*4, we would need to intervene on approximately 1,500 fewer patients than if we were to use SPARRA*v*3 in the same way, in order to achieve the target of avoiding 3,000 admissions (Figure 2F).

SPARRA*v*4 comprises an ensemble of models (see Methods), so we also explored a breakdown of AUROC/AUPRC (Table 2) and calibration (Supplementary Figure S3) across constituent models. The ensemble had slightly better performance than the best constituent models (XGB and RF). Note that some constituent models (ANN, GLM, NB) had ensemble coefficients which were regularised to be vanishingly small, so in practice scores for those models need not be computed when calculating SPARRA*v*4. We investigated whether performance could be improved by using separate sets of coefficients for each SPARRA*v*3 cohort, but found that the improvement was so small that we judged this to be unnecessary (Supplementary Note S3).

**Table 2:**
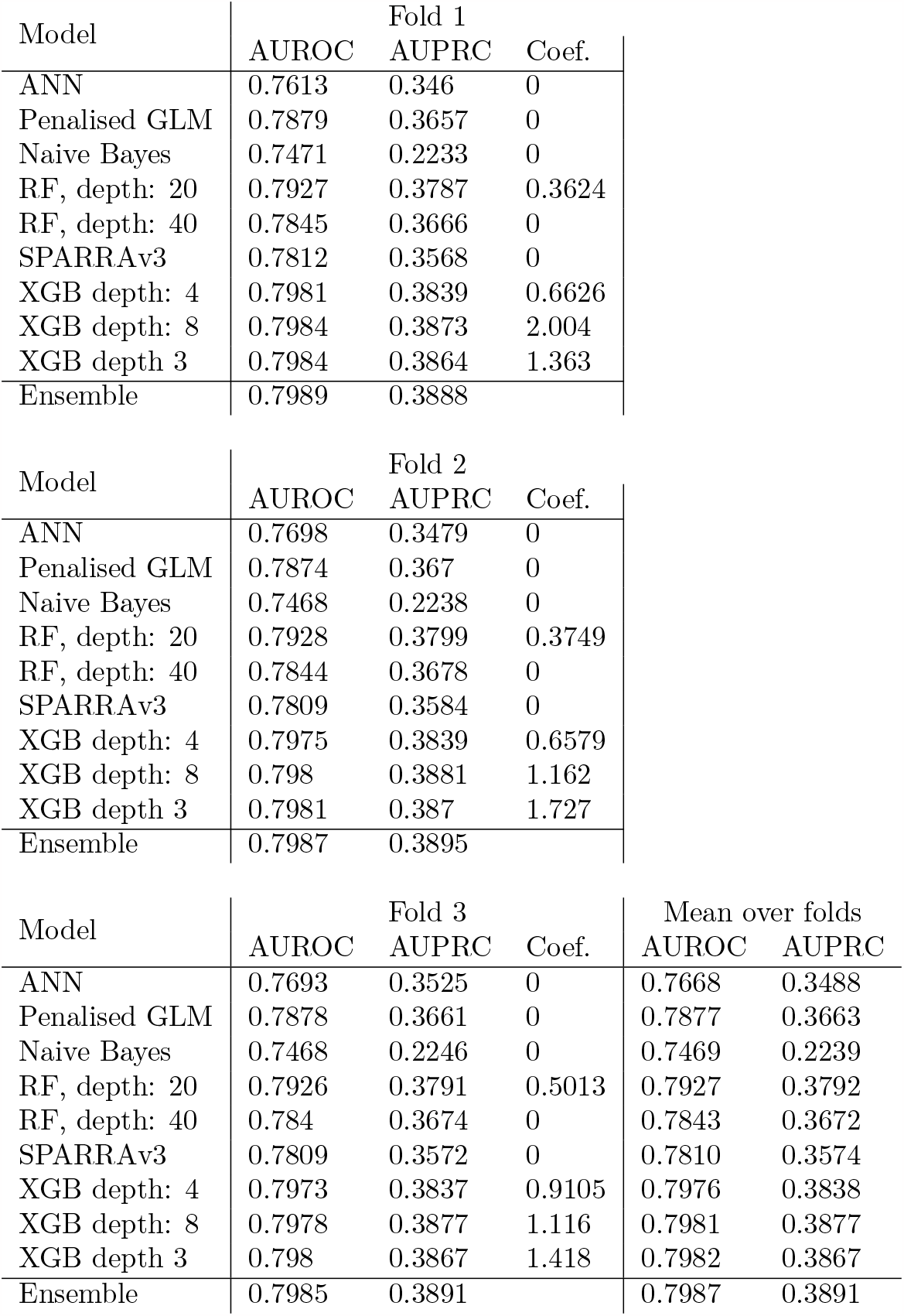
Overall discrimination performance for SPARRAv4 and its constituent models. Areas under ROC curves and PR curves by fold for each constituent predictor and ensemble. Columns ‘Coef.’ indicate estimated coefficients (weights) in the final ensemble (see Methods section for details). All standard errors for AUROCs are *<* 5 × 10^*−*4^ and for AUPRCs are *<* 8 × 10^*−*4^

### Stratified performance of SPARRA*v*3 and SPARRA*v*4

To examine differences in performance more closely, we explored the performance of SPARRA*v*3 and SPARRA*v*4 across different patient subcohorts defined by age, SIMD deciles and the four subcohorts defined as part of SPARRA*v*3 development. Generally, we observed that SPARRA*v*4 had better discrimination performance across all subcohorts (Figure 3A).

**Figure 3:**
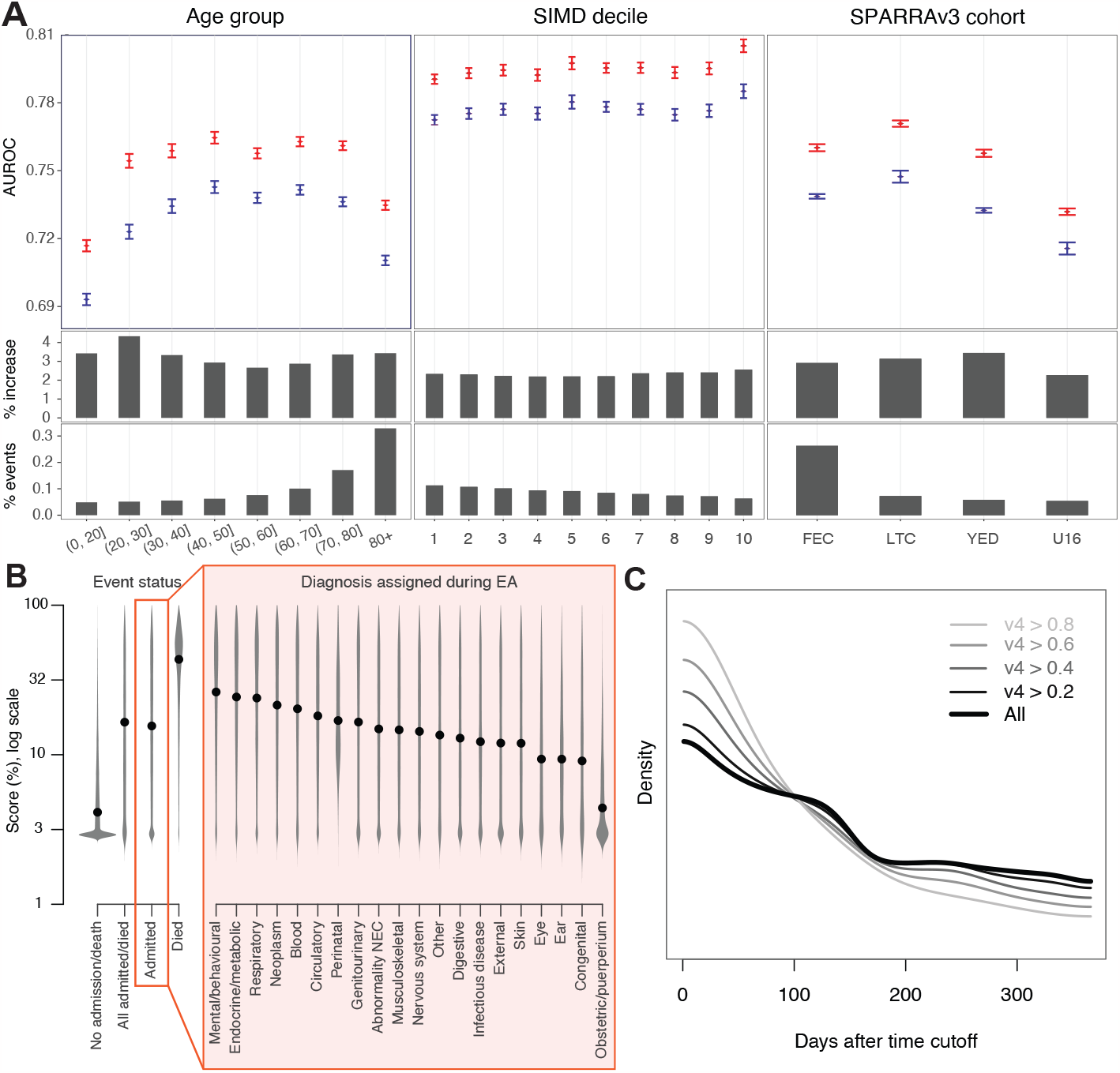
Stratified performance of SPARRA*v*3 and SPARRA*v*4. (A) Performance of SPARRA*v*3 and SPARRA*v*4 in subcohorts defined by age, SIMD and the original subcohorts defined during SPARRA*v*3 development (Methods). Top: AUROC (blue: SPARRA*v*3; red: SPARRA*v*4). Vertical bars denote plus/minus 3 standard deviations. Middle: AUROC increase for SPARRA*v*4 with respect to SPARRA*v*3. For context, bottom sub-panels show the proportion of samples with an event within each group. (B) Distribution of SPARRA*v*4 scores (in log-scale) based on the type of diagnosis recorded during the admission (see Supplementary Table S5 for definitions). Black points indicate the associated medians. Groups were defined according to whether an event was observed (grey violin plots) or, for those with an EA, based on the diagnosis recorded during the admission (black violin plots). (C) Density of time-to-first-EA (that is, days between time cutoff and first EA date) in subsets of individuals who had an EA in the year following the time cutoff and had a SPARRA*v*4 score above a given cutoff. For instance, the lightest line shows density of time-to-first-EA in samples who had an EA and had SPARRA*v*4¿0.8

### Conditional performance of SPARRA*v*4 by admission type and imminence

Figure 3B displays the distribution of SPARRA*v*4 scores stratified according to event status and, for those with an EA, according to the diagnosis that was assigned to the patient during admission (Supplementary Table S5). When comparing samples with and without an event (defined by the composite EA or death outcome), we observed the former had generally lower SPARRA*v*4 scores. Amongst those with an event, all-cause mortality was associated with high SPARRA*v*4 scores. If the event was an EA, we found that samples with certain medical classes of admission tended to have particularly high SPARRA scores, suggesting that such admissions can be predicted disproportionately well (Figure 3B): in particular, those with mental/behavioural, respiratory and endocrine/metabolic related admissions. As one would expect, we were less able to predict external causes of admissions (e.g., S21: open wound of thorax [World Health Organization, 2004]). Obstetric and puerperium-related admissions were particularly challenging to predict by SPARRA*v*4. Similarly, all cause mortality was also associated with high SPARRA*v*4 scores. As one would expect, we were less able to predict external causes of admissions (e.g., S21: open wound of thorax [World Health Organization, 2004]). Obstetric and puerperium-related admissions were particularly challenging to predict by SPARRA*v*4. When further analysing SPARRA*v*4 scores, we also found that among individuals who had an EA during the 1 year outcome period, those with higher risk scores were likelier to have the first EA near the start of the period (Figure 3C). We did not use an absolute threshold to determine who is at high risk. Instead, we ranked individuals according to their scores and looked at those in the top part of the ranking (i.e. with the highest risk scores).

### Deployment scenario stability and performance attenuation

We next addressed two crucial aspects pertaining to practical usage of SPARRA*v*4. Firstly, we assess the durability of performance for a model trained once (at the time cutoff 1 May 2014, using a one-year lookback) and employed to generate scores at future times (1 May and 1 December 2015, 1 May and 1 December 2016, 1 May 2017), confirming it does not deteriorate. This is the way in which SPARRA*v*4 will be deployed by PHS, generating new scores each month but without repeated model updating, akin to SPARRA*v*3’s monthly use without update from 2013–2023. Secondly, we demonstrate that it is none-the-less necessary to update scores despite the absence of model updates, since evolving patient covariates lead to the performance attenuation of any point-in-time score.

We firstly used a *static model M*_0_ (Methods) to predict risk at future time-points (i.e. new scores are generated as the features are updated). *M*_0_ performed essentially equally well over time (Figure 4A-C), with no statistically significant decrease in performance (adjusted p-values *>* 0.05), or improved performance with time for all comparisons of AUROCs. With stability under the deployment scenario confirmed, we also explored the distribution of scores over time. In line with expectations, the quantiles of scores generated by the static model increased as the cohort grew older (Figure 4D). The mean risk scores of individuals in the highest centiles of risk at *t*_0_ decreased over time (Figure 4E), suggesting that very high risk scores tend to be transient. The bivariate densities of time-specific scores (Figure 4F) also show lower scores to be more stable than higher scores, and that subjects ‘jump’ to higher scores (upper left in Figure 4F) more than they drop to lower scores (bottom right).

**Figure 4:**
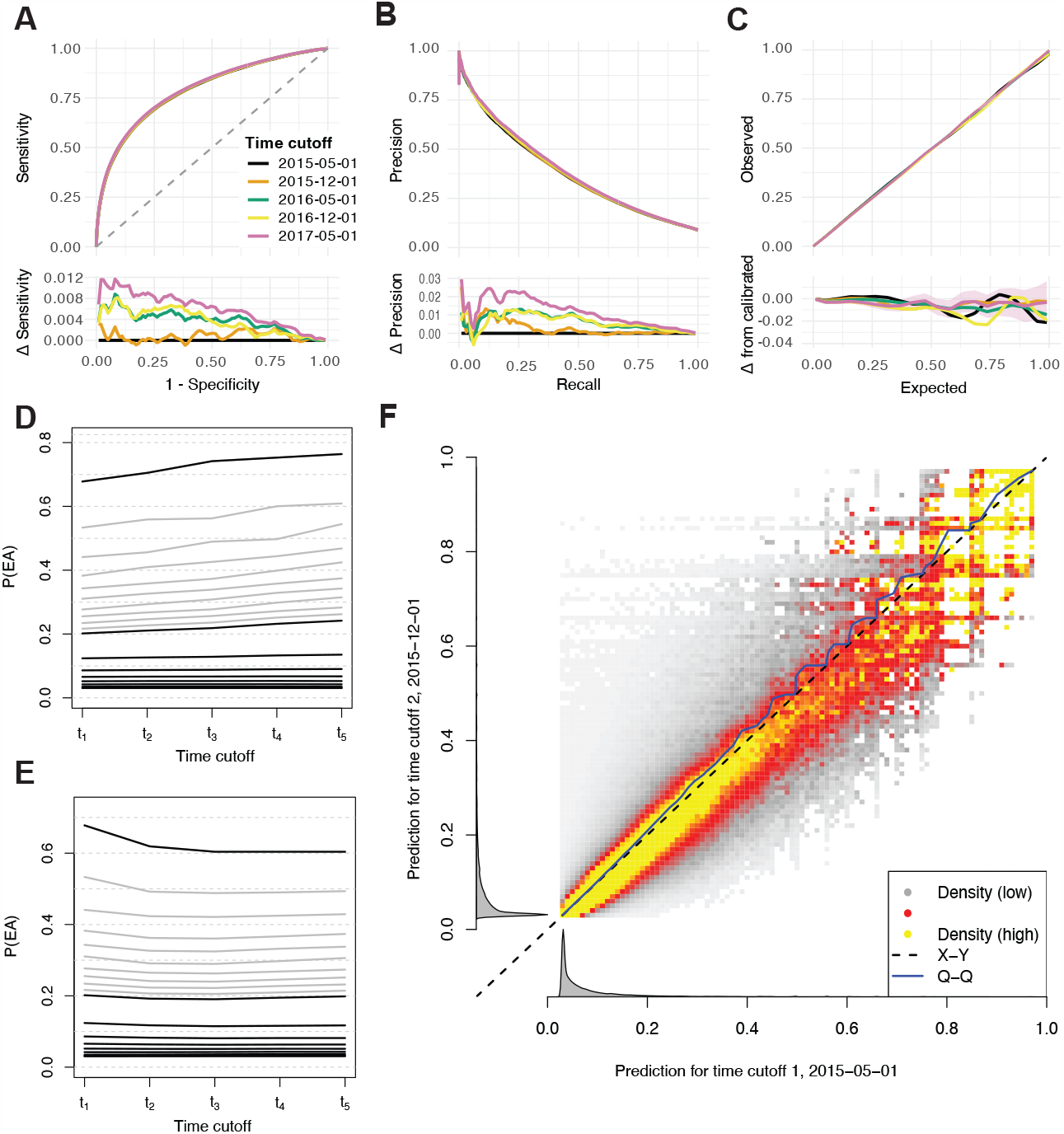
Performance of a static model with changing scores over time. (A-C) Performance of scores calculated at *t*_1_ − *t*_5_ from static model *M*_0_. (A) ROC curves. Lower panel shows differences in sensitivity with respect to *t*_1_. (B) PRC curves. Lower panel shows differences in precision with respect to *t*_1_. (C) Calibration curves. Lower panel shows the difference between observed and expected EA frequency. (D) Centiles (grey) and deciles (black) of risk scores (calculated using *M*_0_) over time, across all individuals with data available at all time cutoffs. (E) Average score over time for groups of individuals defined by risk centiles (grey) and deciles (black) at time *t*_0_ (2 May 2015). (F) Density (low to high: white-grey-red-yellow) of scores generated using the static model *M*_0_ to predict EA risk at *t*_1_ (2 May 2015) and *t*_2_ (1 Dec 2015). The density is normalised to uniform marginal on the Y axis, then the X axis; true marginal distributions of risk scores are shown alongside in grey.

Finally, we examined the behaviour of *static scores* (computed at *t*_0_ using *M*_0_) to predict future event risk (note that the model is also static in this setting, though we will call it *static scores* for brevity). We observed that the static scores performed reasonably well even 2-3 years after *t*_0_, although discrimination and calibration were gradually lost (Supplementary Figure S4A-C). More generally, we observe that scores fitted and calculated at a fixed time cutoff had successively lower AUROCs for predicting EA over future periods (Supplementary Figure S4D). Although the absolute differences in AUROC over time with static scores are small, they are visibly larger than those seen between SPARRA*v*3 and SPARRA*v*4 (Figure 2A), indicating that comparisons analogous to Figure 2E,F would similarly show much larger differences. This affirms the need for updated scores in deployment, despite the static model.

### Feature importance

The features with the largest mean absolute Shapley value (excluding SPARRA*v*3 and the features derived from the topic model) were age, the number of days since the last EA, the number of previous A&E attendances, and the number of antibacterial prescriptions (Table 3). Most features had non-linear effects (see e.g. Supplementary Figure S5A-B). For example, the risk contribution from age was high in infancy, dropping rapidly from infancy through childhood, then remaining stable until around age 65, and rising rapidly thereafter (Figure 5A). We also found a non-linear importance of SIMD (Figure 5B) and number of previous emergency hospital admissions (Supplementary Figure S5C).

**Table 3:** Top 20 most important variables by mean absolute Shapley value (percentage scale). Importance can be interpreted as the average percent added or subtracted to risk score due to this factor.

| Variable | Importance |
| --- | --- |
| Age at time cutoff | 1.530 |
| Days since last emergency admission | 0.752 |
| Number of previous A&E attendances | 0.509 |
| Number of antibacterial prescriptions | 0.376 |
| Number of central nervous system related prescriptions | 0.375 |
| Male sex | 0.373 |
| Days since last A&E attendance | 0.321 |
| SIMD decile | 0.310 |
| Number of emergency bed days | 0.299 |
| Days since last acute admission of any type | 0.285 |
| Days since last outpatient attendance | 0.257 |
| Number of diuretic prescriptions | 0.213 |
| Number of lipid lowering drug prescriptions | 0.194 |
| Number of previous first outpatient appointments | 0.190 |
| Number of recorded long term conditions | 0.173 |
| Number of emergency admissions | 0.161 |
| Total number of filled prescriptions | 0.160 |
| Number of antianaemic prescriptions | 0.159 |
| Number of bronchodilator prescriptions | 0.152 |
| Number of BNF sections from which a prescription was filled | 0.141 |

**Figure 5:**
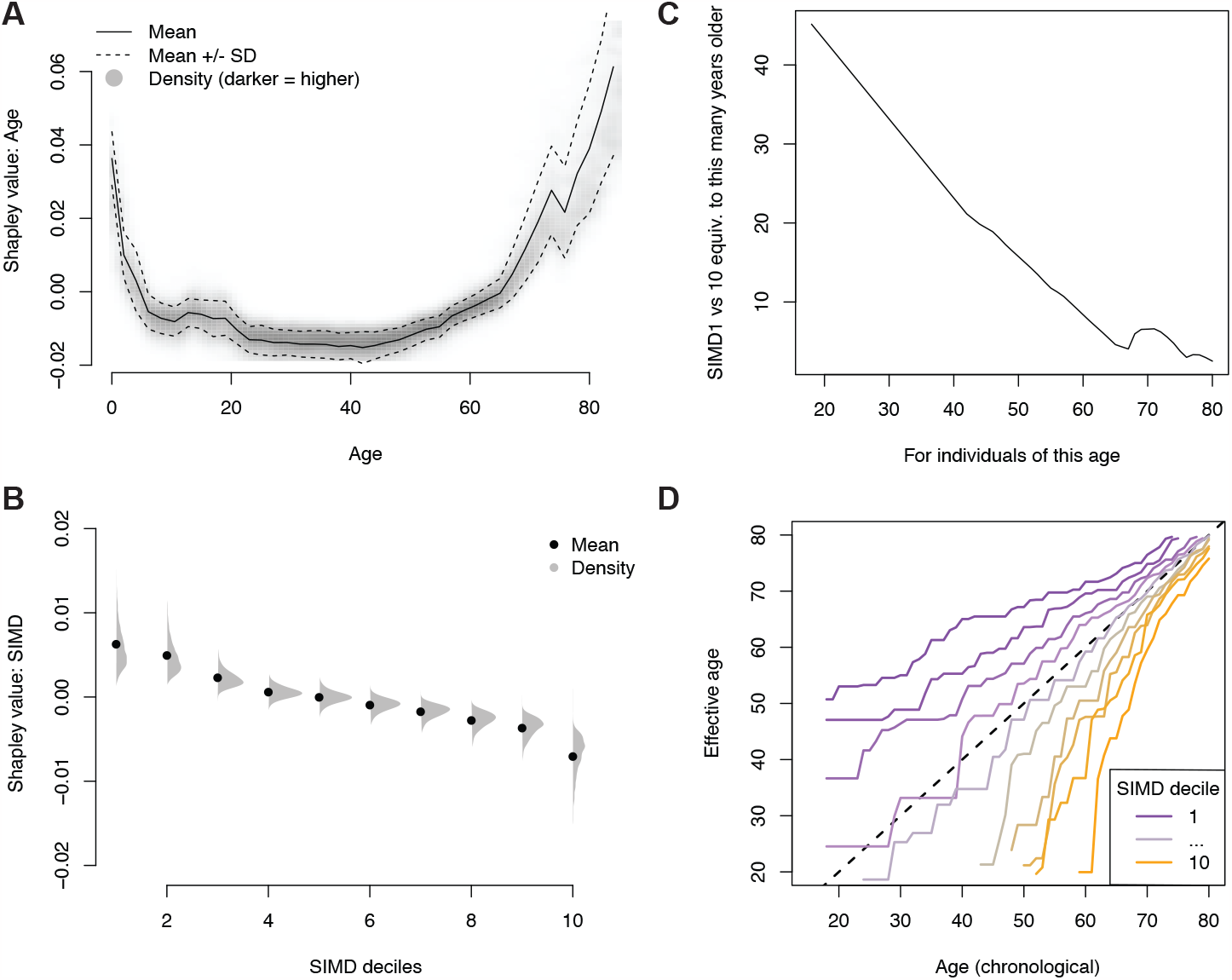
Analysis of Shapley values. Distribution of Shapley values by (A) age and (B) SIMD deciles (1: most deprived; 10: least deprived). (C) Number of additional years of age needed to match the difference in Shapley values between SIMD deciles 1 and 10. (D) ‘Effective ages’ calculated to match EA rates: for an (age, SIMD decile) pair, the age at mean SIMD with the equivalent EA rate.

We further investigated the contribution of SIMD by comparing Shapley values between features. We computed the mean difference in contribution of SIMD to risk score between individuals in the most deprived and least deprived SIMD decile areas, and the additional years of age which would contribute an equivalent amount. This was generally around 10-40 additional years (Figure 5D). In terms of raw admission rates, disparity was further apparent: individuals aged 20 in lowest SIMD decile areas had similar admission rates to individuals aged 70 in the 3 highest SIMD decile areas (Figure 5E).

When exploring the added value (in terms of AUROC) of including the features derived using the topic model (Supplementary Table S4), we observed slightly better performance than the model without such features (*p*-value = 3 × 10^*−*29^; Supplementary Figure S5E-F). In some cases, topic features led to substantial changes in overall score: for example, a topic relating to skin disease contributed more than 2% to the SPARRA*v*4 score (roughly equivalent to the mean contribution to the score from age for individuals aged 75; see Figure 5A) for around 0.43% of individuals with the resultant SPARRA*v*4 scores better-calibrated than the SPARRA*v*3 scores, which did not use a topic model (Supplementary Note S1). Analogously to Figure 2E, we also computed the additional number of samples correctly identified as having an event amongst the top scores by the two models. Although the absolute difference in AUROC was small, we found that the use of topic features increased the number of EAs detected in the top 500,000 scores by around 200.

### Deployment

SPARRA*v*4 was developed in a remote data safe haven (DSH) environment [Public Health Scotland, 2020] without access to internet or modern collaboration tools (e.g. git version control). Whilst our analysis code and a summary of model outputs (e.g. AUROC values) could be securely extracted from the DSH, this was not possible for the actual trained model due to potential leaks of sensitive patient information [Jefferson et al., 2022]. This introduced reproducibility challenges, since the model had to be retrained in a different secure environment before it was deployed by PHS. In particular, this re-development outside the DSH had two distinct phases. Firstly, the raw data transformations (to convert the original databases into a format that is suitable for ML algorithms) were reproduced from scratch from the same source data. Once the output of the transformations matched perfectly between the DSH and the external environment for all features, the topic and predictive models were re-trained. The training process could not be exactly matched due to differing compute environments, package versions and training/validation split. However, after training, the external models were validated by comparing the performance (via AUROC) and the calibration with the results obtained within the DSH.

Another practical issue that arises when developing and deploying a new version of SPARRA is due to potential *performative prediction* effects [Perdomo et al., 2020b]. Since SPARRA*v*3 is already visible to GPs (who may intervene to reduce the risk of high-risk patients), v3 can alter observed risk in training data used for v4, with v3 becoming a *‘victim of its own success’* [Lenert et al., 2019, Sperrin et al., 2019]. *This is potentially hazardous: if some risk factor R* confers high v3 scores prompting GP intervention (e.g., enhanced follow-up), then in the training data for v4, *R* may no longer apparently confer increased risk. Should v4 replace v3, some individuals would therefore have their EA risk underestimated, potentially diverting important anticipatory care away from them. This highlights a critical problem in the theory of model updating [Liley et al., 2021], which we expand on in Supplementary Note and illustrate in Figure 6A-D. As a practical solution, during deployment, GPs could receive the maximum between v3 and v4 scores. This would avoid the potential hazard of risk underestimation, at the cost of mild loss of AUROC (Figure 6E) and score calibration (Figure 6F).

**Figure 6:**
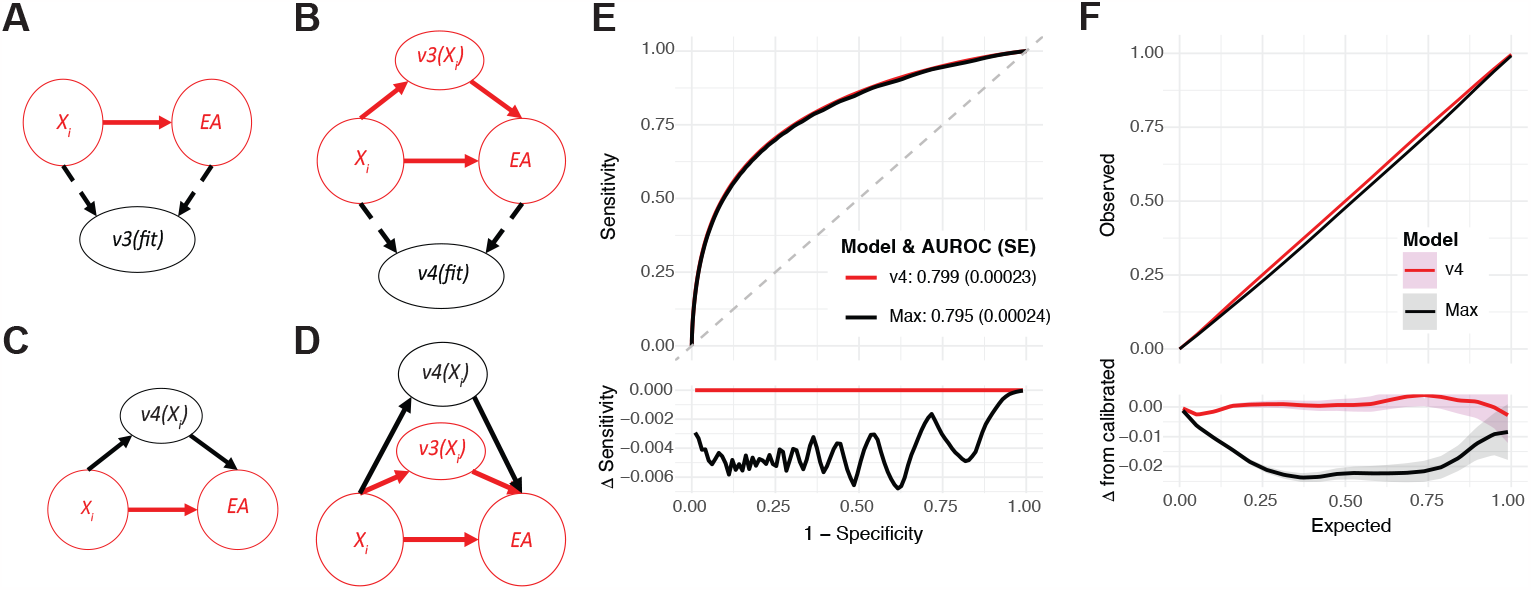
Model updating in the presence of performative effects. (A-D) Causal structure for the training and deployment of SPARRAv3 and SPARRAv4. *X*_*i*_ represents covariates for a patient-time pair; *v*3(*fit*)*/v*4(*fit*) and *v*3(*X*_*i*_)*/v*3(*X*_*i*_) represent the fitting and deployment of v3 and v4 respectively. (A) Training setting for SPARRAv3. (B) Training setting for SPARRAv4. (C) Deployment setting if SPARRAv4 were to naively replace SPARRAv3. (D) Deployment setting in which SPARRAv4 is used as an adjuvant to SPAR-RAv3. (E) Comparison of discrimination (ROC) between SPARRAv4 and the maximum of both scores. (F) Comparison of calibration between SPARRAv4 and the maximum of both scores.

## Discussion

We used routinely collected EHRs from around 5.8 million Scottish residents to develop and evaluate SPARRA*v*4, a risk score that quantifies 1-year EA risk based on age, deprivation (using SIMD as a geographic-based proxy) and a wide range of features derived from a patient’s past medical history. SPARRA*v*4 constitutes a real-world use of ML, derived from population-level data and embedded in clinical settings across Scotland (Figure 1).

While the increases in AUROC and AUPRC over the previous version of SPARRA may be small (Figure 2A,B), the improvement provided by SPARRA*v*4 in terms of absolute benefit to population is substantial (Figure 2E,F). This arises from the use of more flexible ML methods (e.g. to capture non-linear patterns between features and EA risk) and the incorporation of features derived by a topic model which extracts more granular information (with respect to the manually curated features used by SPARRA*v*3) from past diagnoses and prescriptions data. The latter can be thought of as a proxy for multi-morbidity patterns, in that topic models identify patterns of diagnoses and prescriptions which commonly occur together [Kremer et al., 2022], which can be seen to occur in our data (Supplementary Table S4). The use of an ensemble of models also allows stronger models and methods to dominate the final predictor, and weaker models to be discarded.

Our analysis also provides insights into the epidemiology of EA risk, highlighting predictable patterns in terms of EA type (as defined by the recorded primary diagnosis; Figure 3B) and the imminence of EA (Figure 3C), in that those at high risk of an admission are likely to have an imminent admission rather than equally likely to have an admission over the year-long prediction period. Moreover, we studied the contribution of each feature, revealing a complex relationship between age, deprivation and EA risk (Figure 5). Note, however, that we cannot assign a causal interpretation for any reported associations. In particular, the link between SIMD and EA risk is complex; SIMD includes a ‘health’ constituent [Scottish Government, 2016], and individuals in more-deprived SIMD decile areas (1: most deprived; 10: least deprived) miss more primary care appointments [Ellis et al., 2017].

One important strength of SPARRA*v*4 is its nationwide coverage, using existing healthcare databases without the need for additional bespoke data collection. This, however, prevents the use of primary care data (beyond community prescribing) as it is not currently centrally collected in Scotland. Due to privacy considerations, we were also unable to access geographic location data, precluding the study of potential differences between e.g. rural and urban areas and the use of a geographically separated test set [Wallace et al., 2014]. Limited data availability also limits a straightforward comparison of predictive performance (e.g. in terms of AUROC) with respect to similar models developed in England [Billings et al., 2006, Rahimian et al., 2018] (this is also complicated because of different model choices, e.g. [Rahimian et al., 2018] modelled time-to-event data but we used a binary 1-year EA indicator). For example, we do not have information about marital and smoking status, blood test results and family histories; all of which were found to be predictive of EA risk by [Rahimian et al., 2018]. Our training dataset is non-representative of our raw dataset (which in turn is non-representative of the Scottish population, as per Table 1, as is typical of studies based on electronic health records [Verheij et al., 2018, Agniel et al., 2018]), but it does generally include individuals at higher EA risk.

Beyond model development and evaluation, our work also highlights broader challenges that arise in this type of translational project using EHR. In particular, as SPARRA*v*4 has the potential to influence patient care, we have placed high emphasis on transparency and reproducibility while ensuring compliance with data governance constraints. Providing our code in a publicly available repository will also allow us to transparently document future changes to the model (e.g. if any unwanted behaviour is identified during the early stages of deployment). SPARRA*v*4 also constitutes a real-world example in which potential performative effects need to be taken into account when updating an already deployed risk prediction model (Figure 6).

It is critical to emphasise that SPARRA*v*4 will not replace clinical judgement, nor does it direct changes to patient management made solely based on the score. Indeed, any potential interventions must be decided jointly by medical professionals and patients, balancing the underlying risks and benefits. Moreover, lowering EA risk does not necessarily entail overall patient benefit as e.g. long-term oral corticosteroid use in mild asthmatics would reduce EA risk, but the corticosteroids themselves cause an unacceptable cost of long-term morbidity [NICE guidelines, 2017].

Optimal translation into clinical action is a vital research area and is essential for quantifying the benefit of such scores in clinical practice. Indeed, any benefit is dependent on widespread uptake and the existence of timely integrated health and social care interventions, and identification of EA risk is only the first step in this pathway. As such, the evaluation of real-world effectiveness for SPARRA*v*4 and similar risk scores is complex, and requires a multi-disciplinary approach that considers a variety of factors (e.g. the local health economy and the capacity to deliver pre-emtive interventions in primary care) Therefore, we will continue to collaborate to achieve successful deployment of SPARRA*v*4 and will carefully consider the feedback from GPs to improve the model and the communication of its results further (e.g. via informative dashboards). As the COVID-19 pandemic resolves, it will also be important to assess potential effects of dataset shift [Subbaswamy and Saria, 2020] due to disproportionate mortality burden in older individuals and long-term consequences of COVID-19 infections. In an era where healthcare systems are under high stress, we hope that the availability of robust and reproducible risk scores such as SPARRA*v*4 (and its future developments) will contribute to the design of proactive interventions that reduce pressures on healthcare systems and improve healthy life expectancy.

## Methods

### SPARRA*v*3

SPARRA*v*3 [Health and Social Care Information Programme, 2011], deployed in 2012, uses separate logistic regressions on four subcohorts of individuals: frail elderly conditions (FEC; individuals aged *>* 75); long-term conditions (LTC; individuals aged 16-75 with prior healthcare system contact), young emergency department (YED; individuals aged 16-55 who have had at least one A&E attendance in the previous year) and under-16 (U16; individuals aged *<* 16). If an individual belongs to more than one of these groups, the maximum of the associated scores is reported. SPARRA*v*3 was fitted once (at its inception in 2012) with regression coefficients remaining fixed thereafter. Most input features were manually dichotomised into two or more ranges for fitting and prediction. The prediction target for SPARRA*v*3 is EA within 12 months. People who died in the pre-prediction period, and who therefore do not have an outcome for use in the analysis, are excluded. PHS calculated SPARRA*v*3 scores and provided them as input for the analysis described herein. Any GP in Scotland can access SPARRA scores after attaining information governance approval.

### Exclusion criteria

The exclusion criteria were applied per sample (defined as individual-time pairs; Figure 1C). Samples were excluded if: (i) they were excluded from SPARRA*v*3 (these are individuals for which PHS did not calculate a SPARRA*v*3 score and largely correspond to individuals with no healthcare interactions or that were not covered by the four SPARRA*v*3 subcohorts; [Health and Social Care Information Programme, 2011]), (ii) when the individual died prior to the prediction time cutoff, (iii) when the SIMD for the individual was unknown, or (iv) those associated to individuals whose Community Health Index [CHI; ISD Scotland Data Dictionary, 2023] changed during the study period (‘Unmatched’ in Figure 1). The CHI number is a unique identifier which is used in Scotland for health care purposes. Rates of EA and death in the follow-up period were generally lower in excluded samples than in included samples (3.40% versus 8.88%, only considering exclusions which were not due to the individual having died prior to the time cutoff; Supplementary Table S6). Exclusion criteria (i) and (ii) were applied at the sample level, while exclusion criteria (iii) and (iv) were applied at the individual level.

### Feature engineering

A typical entry in the source EHR tables (Supplementary Table S2) recorded a single interaction between a patient and NHS Scotland (e.g. hospitalisation), comprising a unique individual identifier (an anonymised version of the CHI number), the date on which the interaction began (admission), the date it ended (discharge), and further details (diagnoses made, procedures performed). For each sample, entries from up to three years before the time cutoff were considered when building input features, except long-term condition (LTC) records, which considered all data since recording began in January 1981. A full feature list is described in Supplementary Table S3. This includes SPARRA*v*3 [Health and Social Care Information Programme, 2011] features, e.g. age, sex, SIMD deciles and counts of previous admissions (e.g. A&E admissions, drug-and-alcohol-related admissions). Additional features encoding time-since-last-event (e.g. days since last outpatient attendance) were included following findings in [Rahimian et al., 2018]. From community prescribing data, we derived predictors encoding the number of prescriptions of various categories (e.g. respiratory), extending the set of predictors beyond a similar set used in SPARRA*v*3. Similarly to SPARRA*v*3, we also derived the total number of different prescription categories, the total number of filled prescription items, and the number of British National Formulary (BNF) sections from which a prescription was filled [Prasad, 1994]. From LTC records, we extracted the number of years since diagnosis of each LTC (e.g. asthma), the total number of LTCs recorded, and the number of LTCs resulting in hospital admissions.

Data from prescription records and recorded diagnoses tend to be sparse, in that most medications and diagnoses will only be recorded for a small proportion of the population. We used our topic model [Blei et al., 2003] to assimilate this data, by jointly modelling prescriptions and diagnoses using 30 topics (effectively clusters of prescriptions and diagnoses), considering samples as ‘documents’ and diagnoses/prescriptions as ‘words’. This enabled a substantial reduction in feature dimensionality, given the number of diagnoses/prescription factor levels. Using the map from documents to topic probabilities, we used derived topic probabilities as additional features in SPARRA*v*4, which corresponded to sample-wise membership of each topic.

### Choice of prediction target for SPARRAv4

The primary target for SPARRA is to predict whether an individual will experience an EA within 12 months from the prediction cutoff. A problem arises due to the deaths during the follow-up year for which the target may be unknown (e.g. if someone died within 6 months, without a prior EA). Broadly, there are four options for how to treat such individuals during model training and testing:

1. Exclude them from the dataset
2. Treat them according to whether they had an emergency admission before they died
3. Treat them as no admission, or
4. Treat them as an admission

It would also be possible to code death in follow-up differentially; for instance, coding in-hospital death as EA and in-community death as exclusions or non-EA. Our choice not to code all deaths identically is in the interests of non-maleficence. If an individual is at risk of imminent death in the community they will typically be admitted to hospital if it is possible to react in time, with a possible exemption if this is not in their best interests.

Option 1 would exclude the most critically ill individuals from the dataset and hence was discarded. Option 2 would effectively mean such individuals have a follow-up time less than a year, and would classify individuals who died without a hospital admission as having had a ‘desirable’ outcome. Option 3 would effectively classify death as a ‘desirable’ outcome, so we avoided it. The consequences from coding community deaths as non-EA would be severe, as it could mean that healthier individuals at risk of sudden death are either coded as non-EA or excluded from the dataset, potentially leading to inappropriately low scores being assigned to these individuals. This could draw treatment away from individuals in high need. Instead, option 4 allows the general description of the target as ‘a catastrophic breakdown in health’. In this case, our model would not be able to distinguish community deaths from emergency admissions: we may assign high ‘EA’ scores to the very old and terminally ill, when in fact these individuals may be treated in the community rather than admitted. The potential harm from this option is small. It could mean that such individuals are excessively treated rather than palliated, but since palliation over treatment is an active decision [Romo et al., 2017] and such individuals are generally known to be high-risk it is unlikely that the SPARRA score will adversely affect any decisions in this case. As the philosophy of the SPARRA score is to avert breakdowns in health, of which death can be considered an example, we decided to use a composite prediction target (EA or death within 12 months) which is consistent with option 4.

### Machine learning prediction methods

For SPARRA*v*4, we had no prior belief that any ML model class would be best, so considered a range of binary prediction approaches (hereafter referred to as constituent models). The following models were fitted using the *h2o* [LeDell et al., 2019] R package (version 3.24.0.2): an artificial neural network (ANN), two random forests (RF) (depth 20 and 40), an elastic net generalised linear model (GLM) and a naive Bayes (NB) classifier. The xgboost [Chen *et al., 2019*] R package (version 1.6.0.1) was used to train three gradient-boosted trees (XGB) models (maximum tree depth 3, 4, and 8). Hyper-parameter choices are described in Methods. SPARRAv3 was used as an extra constituent model.

Rather than selecting a single constituent model, we used an ensemble approach. Similar to [Van der Laan et al., 2007], we calculated an optimal linear combination (*L*_1_-penalised regression, using the R package glmnet, version 4.1.4) of the scores generated by each constituent model. Ensemble weights were chosen to optimise the AUROC. Finally, we monotonically transformed the derived predictor to improve calibration by inverting the empirical calibration function (Supplementary Note S2).

### Data imputation

As all non-primary care interactions with NHS Scotland are recorded in the input databases, there was no missingness for most features. For ‘time-since-interaction’ type features, samples for which there was no recorded interaction were coded as twice the maximum lookback time. There was minor non-random missingness in topic features (∼ 0.8%) due to individuals in the dataset with no diagnoses or filled prescriptions, for whom topic probabilities could not be calculated. We used mean-value imputation in the ANN and GLM models (deriving mean values from training data only), used missingness to inform tree splits (defaults in [LeDell et al., 2019]) in RF, used sample-wise imputation in XGB (as per [Chen et al., 2019]) and dropped during fitting (default in [LeDell et al., 2019]) in NB (omitted missing values for prediction). All imputation rules were determined using training sets only.

Particular care was required for features encoding total lengths of hospital stays. In some cases, a discharge date was not recorded, which could lead to an erroneous assumption of a very long hospital stay (from admission until the time cutoff). To address this, we truncated apparently spuriously long stays at data-informed values (Supplementary Note S4).

### Hyperparameter choice for ML prediction methods

We used a range of constituent models. The h2o [LeDell et al., 2019] R package (version 3.24.0.2) was used to train ANN, RF, GLM and NB models. The xgboost [Chen et al., 2019] R package (version 1.6.0.1) was used to train the XGB models. Unless otherwise specified, hyperparameters were set as the software defaults. When tuned, hyperparameter values were chosen to optimise the default objective functions implemented for each method: logloss or the ANN, RFs and GLM, likelihood for the NB model; and a logistic objective for the XGB trees. In all cases, hyperparameters were determined by randomly splitting the relevant dataset into a training and test set of 80% and 20% of the data respectively. Details for each method are provided below. Only limited hyperparameter tuning was possible due to the restricted computational environment in the data safe haven (see Results)

### SPARRAv3

SPARRAv3 scores were calculated by PHS using their existing algorithm Health and Social Care Information Programme [2011].

### Artificial neural network (ANN)

We used a training dropout rate of 20% to reduce generalisation error. We optimised over the number of layers (1 or 2) and the number of nodes in each layer (128 or 256).

### Random forest (RF)

We fitted two RF: one had maximum depth 20 and 500 trees, and the other had maximum depth 40 and 50 trees (both taking a similar time to fit).

### Gradient-boosted trees (XGB)

We fitted three boosted tree models with three maximum depths: 3, 4, and 8. For the deeper-tree model, we set a low step size shrinkage *η* = 0.075 and a positive minimum loss reduction *γ* = 5 in order to avoid overfitting. In the other two models, we used default values of *η* = 0.3, *γ* = 0.

### Naive Bayes (NB)

The only hyperparameter we tuned was a Laplace smoothing parameter, varying between 0 and 4.

### Penalised Generalised linear model (GLM)

We optimised *L*_1_ and *L*_2_ penalties (an elastic net), considering total penalty (*L*_1_ + *L*_2_) in 10^*−{*1,2,3,4,5*}*^, and a ratio *L*_1_*/L*_2_ in {0, 0.5, 1}.

### Cross-validation

We fitted and evaluated SPARRA*v*4 using three-fold cross-validation (CV). We considered three-fold cross validation acceptable in our case given the size of our dataset [Bates et al., 2023]. This was designed such that all elements of the model evaluated on a test set were agnostic to samples in that test set. Individuals were randomly partitioned into three data folds (F1, F2 and F3). At each CV iteration, F1 and F2 were combined and used as a training dataset, F3 was used as a test dataset. The training dataset (F1+F2) was used to fit the topic model and to train all constituent models (except SPARRA*v*3, whose training anyhow pre-dates the data used here). The ensemble weights and re-calibration transformation were learned using F1 + F2, i.e. without using the test set from the test set (Supplementary Note S2).

### Predictive performance

Our primary endpoint for model performance was AUROC. We also considered area-underprecision-recall curves (PRC) and calibration curves. We plotted calibration curves using a kernelised calibration estimator (Supplementary Note S5).

For simplicity, figures show ROC/PRC that were calculated by combining all samples from the three *test* CV folds (that is, all scores and observed outcomes were merged to draw a single curve). Quoted AUROC/AUPRC values were calculated as an average across the three *test* CV folds to avert problems from between-fold differences in models [Forman and Scholz, 2010]. For ease of comparison, we also used mean-over-folds to compute quoted AUROCs and AUPRCs for SPARRA*v*3, although the latter was not fitted to our data.

### Deployment scenario stability and performance attenuation

Using the same analysis pipeline as for the development of SPARRA*v*4, we trained a static model *M*_0_ to an early time cutoff (*t*_0_=1 May 2014), and using one year of data prior to *t*_0_ to derive predictors (the restricted lookback is the only deviation from the actual model pipeline, due to limited temporal span of the training data).

We studied the performance of *M*_0_ as a *static model* to repeatedly predict risk at future time cutoffs, which mirrors the way in which PHS will deploy the model. To do this, we assembled test features from data 1 year prior to *t*_1_=1 May 2015, *t*_2_=1 Dec 2015, *t*_3_=1 May 2016, *t*_4_=1 Dec 2016, and *t*_5_=1 May 2017, applying *M*_0_ to predict EA risk in the year following each time-point. In this analysis, the comparison of the distribution of scores over time only considered the cohort of patients who were alive and had valid scores at *t*_1_, …, *t*_5_.

To ensure a fair comparison when evaluating the performance of *static scores* (computed at *t*_0_ using *M*_0_) to predict future event risk (at *t*_1_, …, *t*_5_), we only considered a subsample of 1 million individuals with full data across all time-points, selected such that global admission rates matched those at *t*_0_.

### Assessment of feature importance

We examined the contribution of feature to risk scores at an individual level by estimating Shapley values [Lundberg and Lee, 2017] for each feature. For simplicity, this calculation was done using 20,000 randomly-chosen samples in the first cross-validation fold (F1). We treated SPARRA*v*3 scores as fixed predictors rather than as functions of other predictors.

We also assessed the added value of inclusion of topic-model derived features, which summarise more granular information about the previous medical history of a patient with respect to those included in SPARRA*v*3. For this purpose, we refitted the model to F2+F3 with topic-derived features excluded from the predictor matrix. We compared the performance of these models using F1 as test data. We compared the performance of predictive models with and without the features derived from the topic model by comparing AUROC values using DeLong’s test [DeLong et al., 1988].

### Model updating in the presence of performative effects

We aim to produce the SPARRA score to accurately estimate EA risk over a year under normal medical care. In other words, the score should represent the EA risk if GPs do not already have access to such a risk score. Because GPs see a SPARRA score (SPARRAv3) and may act on it, the observed risk may be lower than predicted - the score may become a ‘victim of its own success’ [Lenert et al., 2019, Sperrin et al., 2019] due to performative effects Perdomo et al. [2020a]. Unfortunately, since the SPARRAv3 score is widely available to Scottish GPs, and may be freely acted on, we cannot assess the behaviour of the medical system in its absence. This is potentially hazardous [Liley et al., 2021].

Formally, at a given fixed time, for each individual, the value of ‘EA in the next 12 months’ is a Bernoulli random variable. The probability of the event for individual *i* is conditional on a set of covariates *X*_*i*_ derived from their EHR. We denote *v*3(*X*_*i*_), *v*4(*X*_*i*_) the derived SPARRAv3 and SPARRAv4 scores as functions of covariates, and assume a causal structure shown in Figure 6 (for simplicity, we assume there are no unobserved confounders but the same argument applies in their presence). With no SPARRA-like predictive score in place, there is only one causal pathway *X*_*i*_ → *EA*. It is to this system (coloured red) that *v*3 was fitted. Here, *v*3(*X*_*i*_) estimates the ‘native’ risk *Pr*(*EA*|*X*_*i*_) (ignoring previous versions of the SPARRA score, which covered *<* 30% of the population). Although *v*3(*X*_*i*_) is determined entirely by *X*_*i*_, the act of distributing values of *v*3(*X*_*i*_) to GPs opens a second causal pathway from *X*_*i*_ to *EA* (Figure 6) driven by GP interventions made in response to *v*3(*X*_*i*_) scores. It is to this system (coloured red) that SPARRAv4 is fitted. Hence, *v*4(*X*_*i*_) is an estimator of *Pr*(*EA* | *X*_*i*_, *v*3(*X*_*i*_)), a ‘conditional’ risk after interventions driven by *v*3(*X*_*i*_) have been implemented.

If SPARRAv4 naively replaced SPARRAv3 (Figure 6), we would be using *v*4(*X*_*i*_) to predict behaviour of a system different to that on which it was trained (Figure 6). To amend this problem, we propose to use SPARRAv4 in *conjunction* with SPARRAv3 rather than to completely replace it (Figure 6). Ideally, GPs would be given *v*3(*X*_*i*_) and *v*4(*X*_*i*_) simultaneously and asked to *firstly* observe and act on *v*3(*X*_*i*_), *then* observe and act on *v*4(*X*_*i*_), thereby only using *v*4(*X*_*i*_) as per Figure 6. This is impractical, so instead, we propose to distribute a single value (given by the maximum between *v*3(*X*_*i*_) and *v*4(*X*_*i*_)), avoiding the potential hazard of risk underestimation, at the cost of mild loss of score calibration (Figure 6).

## Data Availability

Raw data for this project are patient-level NHS Scotland health records, and are confidential. Due to the confidential nature of the data used, all analysis took place on remote 'safe havens', without access to internet, software updates or unpublished software. Information Governance training was required for all researchers accessing the analysis environment. Moreover, to avoid the risk of accidental disclosure of sensitive information, an independent team carried out statistical disclosure control checks to all data exports, including the outputs presented in this manuscript. All analysis code and co-ordinates required to reproduce our Figures are available in github.com/jamesliley/SPARRAv4

## Supporting information

**Supplementary Table S1** Checklist for TRIPOD guidelines [Collins et al., 2015].

**Supplementary Table S2** Definition of input features for SPARRAv4

**Supplementary Table S3** Definition of input features for SPARRA*v*4.

**Supplementary Table S4** Exploration of contributors to each topic.

**Supplementary Table S5** Definition of different admission types.

**Supplementary Table S6** Frequency of admissions and deaths amongst excluded samples.

**Supplementary Table S7** Coefficients of ensemble when fitted separately to SPARRA*v*3 cohorts.

**Supplementary Figure S1** Extended data overview.

**Supplementary Figure S2** Density plot comparing SPARRA*v*3 and SPARRA*v*4 scores.

**Supplementary Figure S3** Calibration curves for SPARRA*v*4 model constituents.

**Supplementary Figure S4** Performance of a static model and static scores used to predict risk at future time cutoffs.

**Supplementary Figure S5** Feature importance

**Supplementary Note S1** Analysis of specific effects of a topic feature.

**Supplementary Note S2** Details of the re-calibration procedure.

**Supplementary Note S3** Investigation of use of SPARRA*v*3-cohort specific ensemble coefficients

**Supplementary Note S4** Imputation of lengths of stay when discharge date was missing.

**Supplementary Note S5** Assessment of calibration.

### Code and data sharing

Raw data for this project are patient-level EHR, and are confidential. Due to the confidential nature of the data, all analysis took place on a remote “data safe haven”, without access to internet, software updates or unpublished software. Information Governance training was required for all researchers accessing the analysis environment. Moreover, to avoid the risk of accidental disclosure of sensitive information, an independent team carried out statistical disclosure control checks on all data exports, including the outputs presented in this manuscript. All analysis code and co-ordinates required to reproduce our Figures are available in github.com/jamesliley/SPARRA*v*4. This manuscript conforms to the TRIPOD guidelines [Collins et al., 2015] (Supplementary Table S1).

## Ethics statement

The project was covered under National Safe Haven Generic Ethical Approval (favourable ethical opinion from the East of Scotland NHS Research Ethics Service).

## Acknowledgements

The authors note that this project’s success was entirely contingent on close co-operation between the Alan Turing Institute and PHS. We thank all individuals involved in primary care in Scotland for the continued support of the SPARRA project and the Public Benefit and Privacy Panel for Health and Social Care (study number 1718-0370) for Information Governance approval on behalf of the Health Boards in NHS Scotland.

All author contributions were significant and essential to the completion of this work. Author contributions were as follows: Manuscript preparation: JL, SRE, BAM, SJV, CAV, LJMA, IT; Project initiation: SJV, CAV, LJMA, CH; Model design: JL, GB, SJV, CAV, LJMA; Code and scripts: JL, GB, LJMA; NC; IT; SDR; Code review and checking: SRE, IT; SDR; Setup of computational system: GB, LJMA; Data access management: DC, RP; EHR access: KB, DC, JI, RP, SO, SR; Public health input: KB, DC, SO, JI, RP, SR; Medical input: JL, BAM, KM; Core planning group: JL, GB, SRE, BAM, KB, DC, JI, KM, RP, SJV, CAV, LJMA; Logistical and legal oversight of project: SH, KP.

Computing for this project was performed in the Scottish National Safe Haven (NSH), which is commissioned by eDRIS, Public Health Scotland from EPCC, based at The University of Edinburgh. The authors would like to acknowledge the support of the eDRIS Team for their involvement in obtaining approvals, provisioning and linking data and the use of the secure analytical platform within the National Safe Haven.

We thank the Alan Turing Institute, PHS, the MRC Human Genetics Unit at the University of Edinburgh, Durham University, University of Warwick, Wellcome Trust, Health Data Research UK, and King’s College Hospital, London for their continuous support of the authors. JL, IT, CAV and LJMA were partially supported by Wave 1 of The UKRI Strategic Priorities Fund under the EPSRC Grant EP/T001569/1, particularly the “Health” theme within that grant and The Alan Turing Institute; JL, IT, BAM, CAV, LJMA and SJV were partially supported by Health Data Research UK, an initiative funded by UK Research and Innovation, Department of Health and Social Care (England), the devolved administrations, and leading medical research charities; SJV, NC and GB were partially supported by the University of Warwick Impact Fund. SRE is funded by the EPSRC doctoral training partnership (DTP) at Durham University, grant reference EP/R513039/1; LJMA was partially supported by a Health Programme Fellowship at The Alan Turing Institute; CAV was supported by a Chancellor’s Fellowship provided by the University of Edinburgh.

For the purpose of open access, the author has applied a Creative Commons Attribution (CC BY) licence to any Author Accepted Manuscript version arising from this submission.

## SUPPLEMENTARY TABLES

**Table S1.**
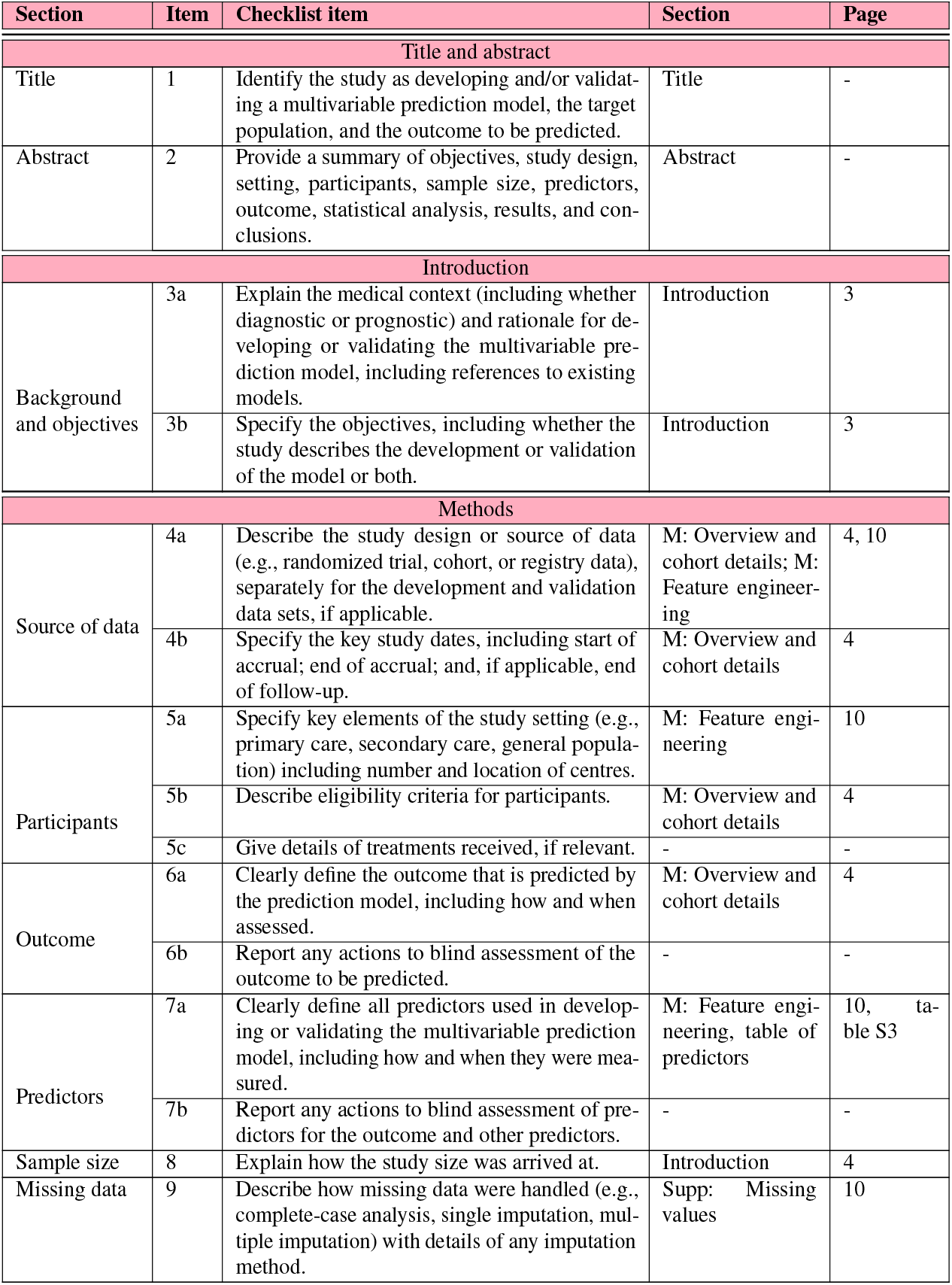
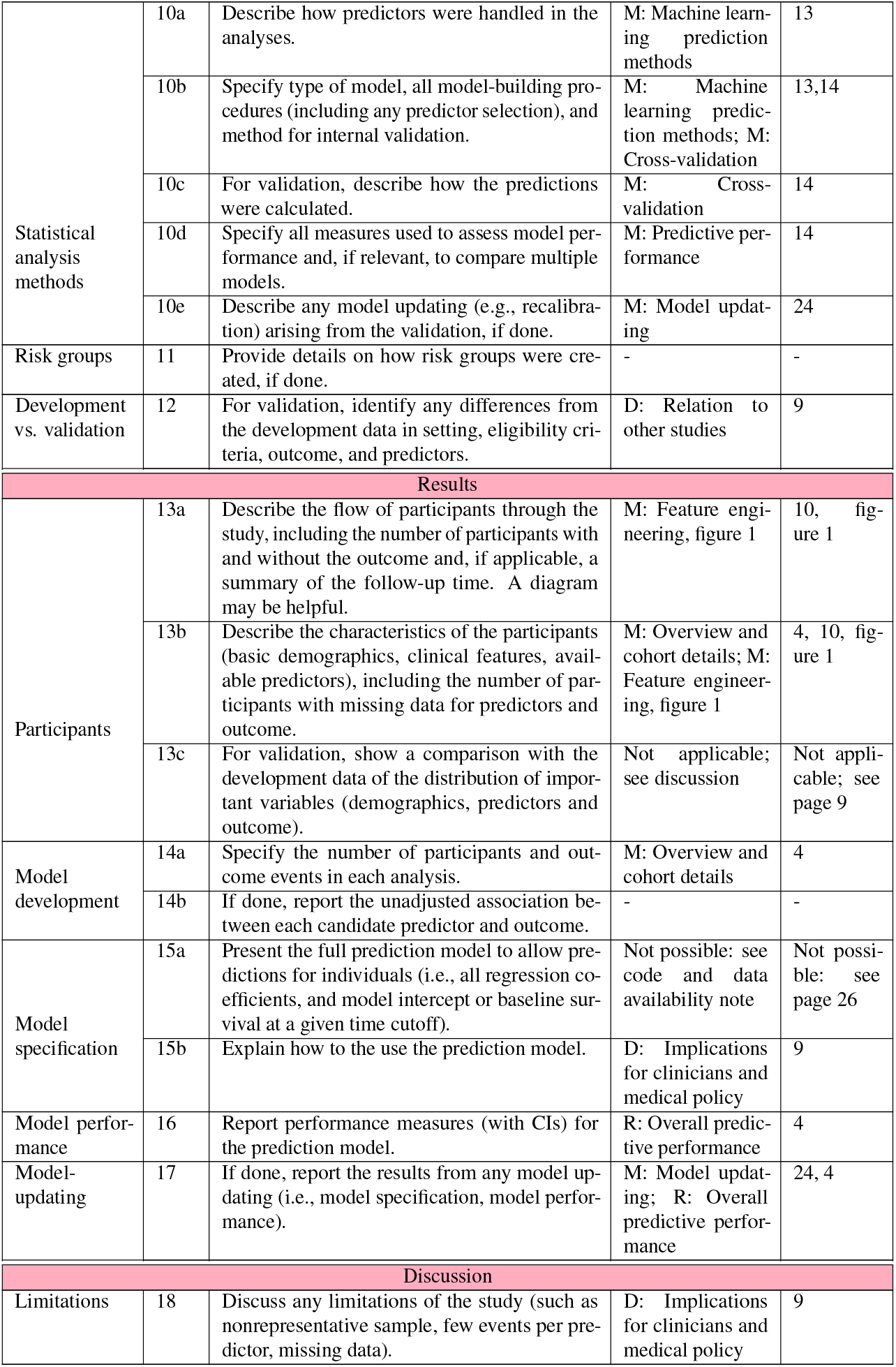
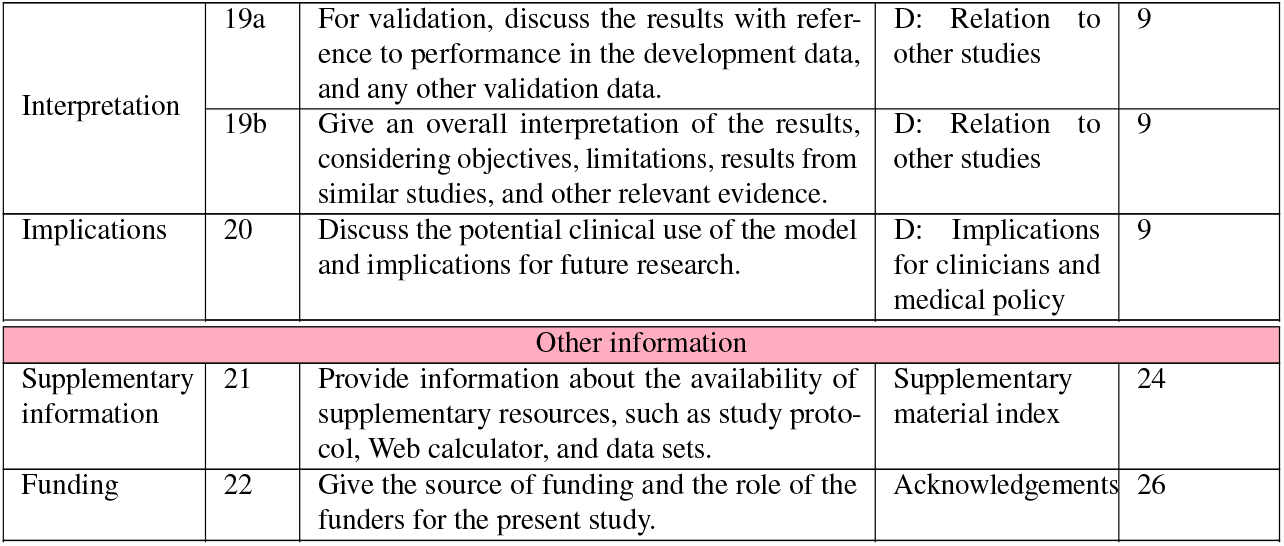
TRIPOD guidelines and pages where discussed [Moons et al., 2015]

| Section | Item | Checklist item | Section | Page |
| --- | --- | --- | --- | --- |
| Title and abstract |  |  |  |  |
| Title | 1 | Identify the study as developing and/or validating a multivariable prediction model, the target population, and the outcome to be predicted. | Title | - |
| Abstract | 2 | Provide a summary of objectives, study design, setting, participants, sample size, predictors, outcome, statistical analysis, results, and conclusions. | Abstract | - |
| Introduction |  |  |  |  |
| Background and objectives | 3a | Explain the medical context (including whether diagnostic or prognostic) and rationale for developing or validating the multivariable prediction model, including references to existing models. | Introduction | 3 |
|  | 3b | Specify the objectives, including whether the study describes the development or validation of the model or both. | Introduction | 3 |
| Methods |  |  |  |  |
| Source of data | 4a | Describe the study design or source of data (e.g., randomized trial, cohort, or registry data), separately for the development and validation data sets, if applicable. | M: Overview and cohort details; M: Feature engineering | 4, 10 |
|  | 4b | Specify the key study dates, including start of accrual; end of accrual; and, if applicable, end of follow-up. | M: Overview and cohort details | 4 |
| Participants | 5a | Specify key elements of the study setting (e.g., primary care, secondary care, general population) including number and location of centres. | M: Feature engineering | 10 |
|  | 5b | Describe eligibility criteria for participants. | M: Overview and cohort details | 4 |
|  | 5c | Give details of treatments received, if relevant. | - | - |
| Outcome | 6a | Clearly define the outcome that is predicted by the prediction model, including how and when assessed. | M: Overview and cohort details | 4 |
|  | 6b | Report any actions to blind assessment of the outcome to be predicted. | - | - |
| Predictors | 7a | Clearly define all predictors used in developing or validating the multivariable prediction model, including how and when they were measured. | M: Feature engineering, table of predictors | 10, table S3 |
|  | 7b | Report any actions to blind assessment of predictors for the outcome and other predictors. | - | - |
| Sample size | 8 | Explain how the study size was arrived at. | Introduction | 4 |
| Missing data | 9 | Describe how missing data were handled (e.g., complete-case analysis, single imputation, multiple imputation) with details of any imputation method. | Supp: Missing values | 10 |
| Statistical analysis methods | 10a | Describe how predictors were handled in the analyses. | M: Machine learning prediction methods | 13 |
|  | 10b | Specify type of model, all model-building procedures (including any predictor selection), and method for internal validation. | M: Machine learning prediction methods; M: Cross-validation | 13,14 |
|  | 10c | For validation, describe how the predictions were calculated. | M: Cross-validation | 14 |
|  | 10d | Specify all measures used to assess model performance and, if relevant, to compare multiple models. | M: Predictive performance | 14 |
|  | 10e | Describe any model updating (e.g., recalibration) arising from the validation, if done. | M: Model updating | 24 |
| Risk groups | 11 | Provide details on how risk groups were created, if done. | - | - |
| Development vs. validation | 12 | For validation, identify any differences from the development data in setting, eligibility criteria, outcome, and predictors. | D: Relation to other studies | 9 |
| Results |  |  |  |  |
| Participants | 13a | Describe the flow of participants through the study, including the number of participants with and without the outcome and, if applicable, a summary of the follow-up time. A diagram may be helpful. | M: Feature engineering, figure 1 | 10, figure 1 |
|  | 13b | Describe the characteristics of the participants (basic demographics, clinical features, available predictors), including the number of participants with missing data for predictors and outcome. | M: Overview and cohort details; M: Feature engineering, figure 1 | 4, 10, figure 1 |
|  | 13c | For validation, show a comparison with the development data of the distribution of important variables (demographics, predictors and outcome). | Not applicable; see discussion | Not applicable; see page 9 |
| Model development | 14a | Specify the number of participants and outcome events in each analysis. | M: Overview and cohort details | 4 |
|  | 14b | If done, report the unadjusted association between each candidate predictor and outcome. | - | - |
| Model specification | 15a | Present the full prediction model to allow predictions for individuals (i.e., all regression coefficients, and model intercept or baseline survival at a given time cutoff). | Not possible: see code and data availability note | Not possible: see page 26 |
|  | 15b | Explain how to use the prediction model. | D: Implications for clinicians and medical policy | 9 |
| Model performance | 16 | Report performance measures (with CIs) for the prediction model. | R: Overall predictive performance | 4 |
| Model-updating | 17 | If done, report the results from any model updating (i.e., model specification, model performance). | M: Model updating; R: Overall predictive performance | 24, 4 |
| Discussion |  |  |  |  |
| Limitations | 18 | Discuss any limitations of the study (such as nonrepresentative sample, few events per predictor, missing data). | D: Implications for clinicians and medical policy | 9 |
| Interpretation | 19a | For validation, discuss the results with reference to performance in the development data, and any other validation data. | D: Relation to other studies | 9 |
|  | 19b | Give an overall interpretation of the results, considering objectives, limitations, results from similar studies, and other relevant evidence. | D: Relation to other studies | 9 |
| Implications | 20 | Discuss the potential clinical use of the model and implications for future research. | D: Implications for clinicians and medical policy | 9 |
| Other information |  |  |  |  |
| Supplementary information | 21 | Provide information about the availability of supplementary resources, such as study protocol, Web calculator, and data sets. | Supplementary material index | 24 |
| Funding | 22 | Give the source of funding and the role of the funders for the present study. | Acknowledgements | 26 |

**Table S2.** Input data sources. The data comprises national EHR Scottish databases between 1 May 2013 and 30 April 2018. Each record corresponds to a single interaction with the health system. SMR - Scottish Morbidity Records. Information about specific EHR tables is available at Public Health Scotland [2023] (SMR datasets), Public Health Scotland [2020a] (A&E2), Public Health Scotland [2020c] (System Watch) and Public Health Scotland [2020b] (PIS). The LTC table was derived by PHS from historic SMR01 tables with an admission between 01 January 1981 and 30 April 2018. The Deaths table was provided to PHS by National Records of Scotland (NRS) and is up to date until TBC. The timescale for the pre-prediction period is 3 years for PIS, SMR00, SMR01, System Watch, A&E2, SMR01E and SMR04. All LTC records up to 01 January 1981 are used in the pre-prediction period. The earlier end of the pre-prediction period is because the most recent month will not be generally available when running the predictions.

| Type | Name | Description | Records | Individuals | End of pre-prediction (before start of risk year) |
| --- | --- | --- | --- | --- | --- |
| Raw EHR | PIS | GP prescribing information | 393,573,549 | 5,589,772 | One month based upon the Paid Date |
|  | SMR00 | Outpatient attendances | 27,463,987 | 3,753,240 | One day based upon the Clinic Date |
|  | SMR01 | Acute inpatients and day cases | 26,326,889 | 2,205,606 | One day based upon the Date of Admission |
|  | SystemWatch | Urgent care monitoring | 12,890,591 | 1,750,009 | One day based upon the Date of Admission |
|  | A&E2 | Accident and emergency records | 7,539,454 | 3,031,773 | One day based upon the Date of Arrival |
|  | SMR01E | Geriatric long stay | 137,340 | 25,015 | . |
|  | SMR04 | Mental health inpatient and day cases | 111,487 | 51,635 | One day based upon the Date of Admission |
|  | **All raw EHR tables combined** |  | 468,043,297 | 5,829,532 |  |
| Other | SPARRALTC | Long-term conditions | 3,286,987 | 1,978,171 | . |
|  | Deaths | Mortality records | 283,554 | 283,554 |  |

**Table S3.**
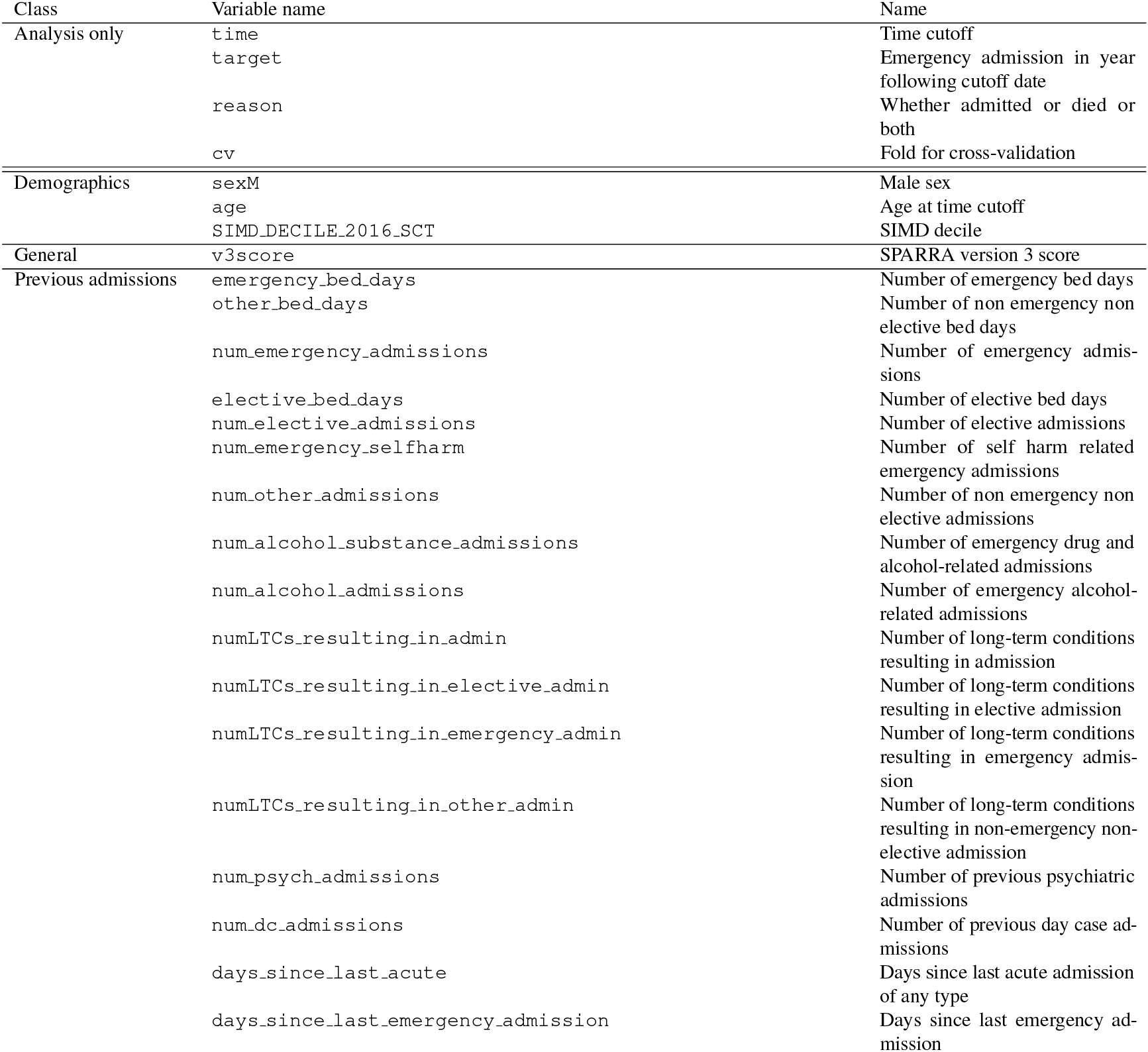
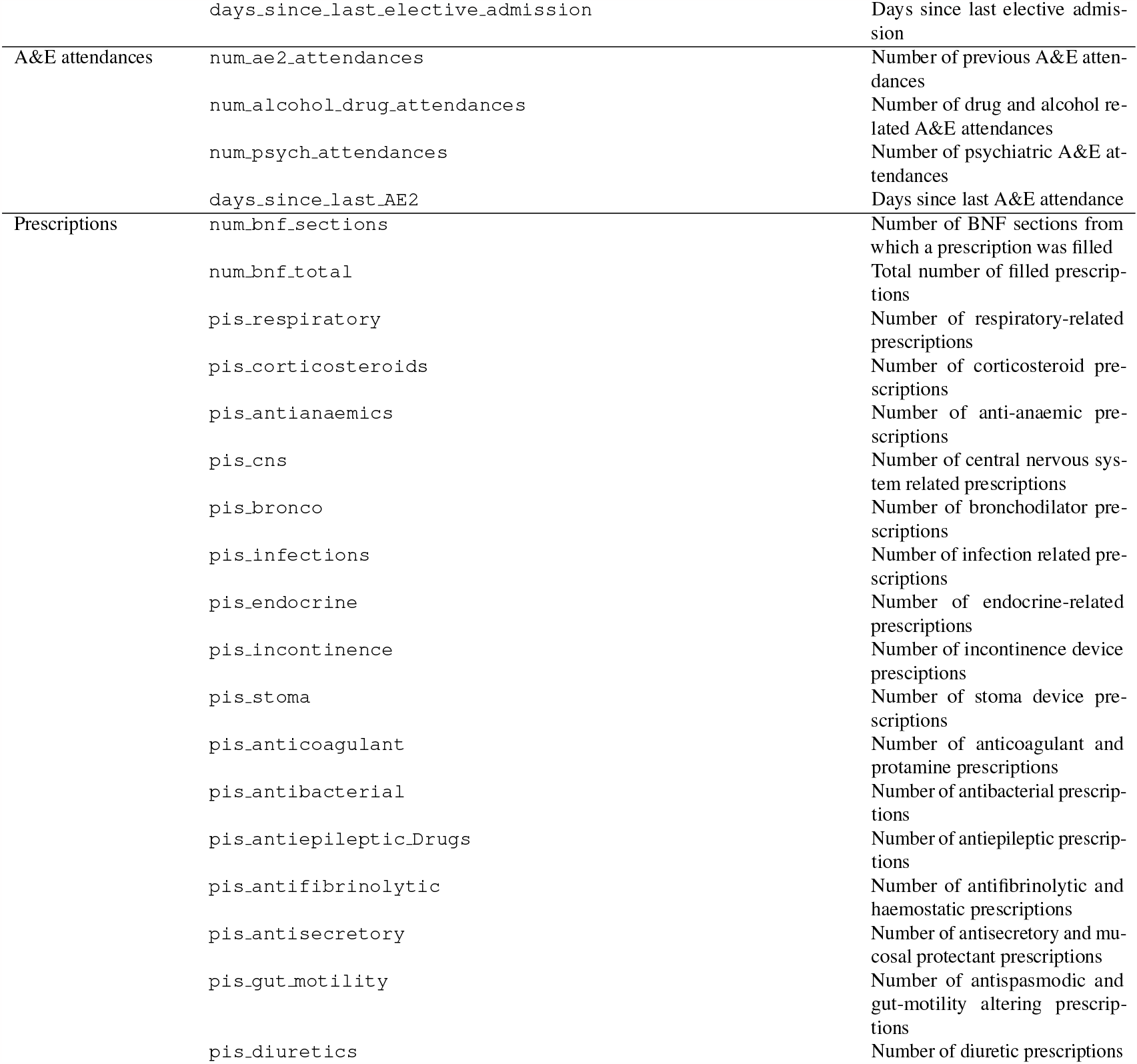
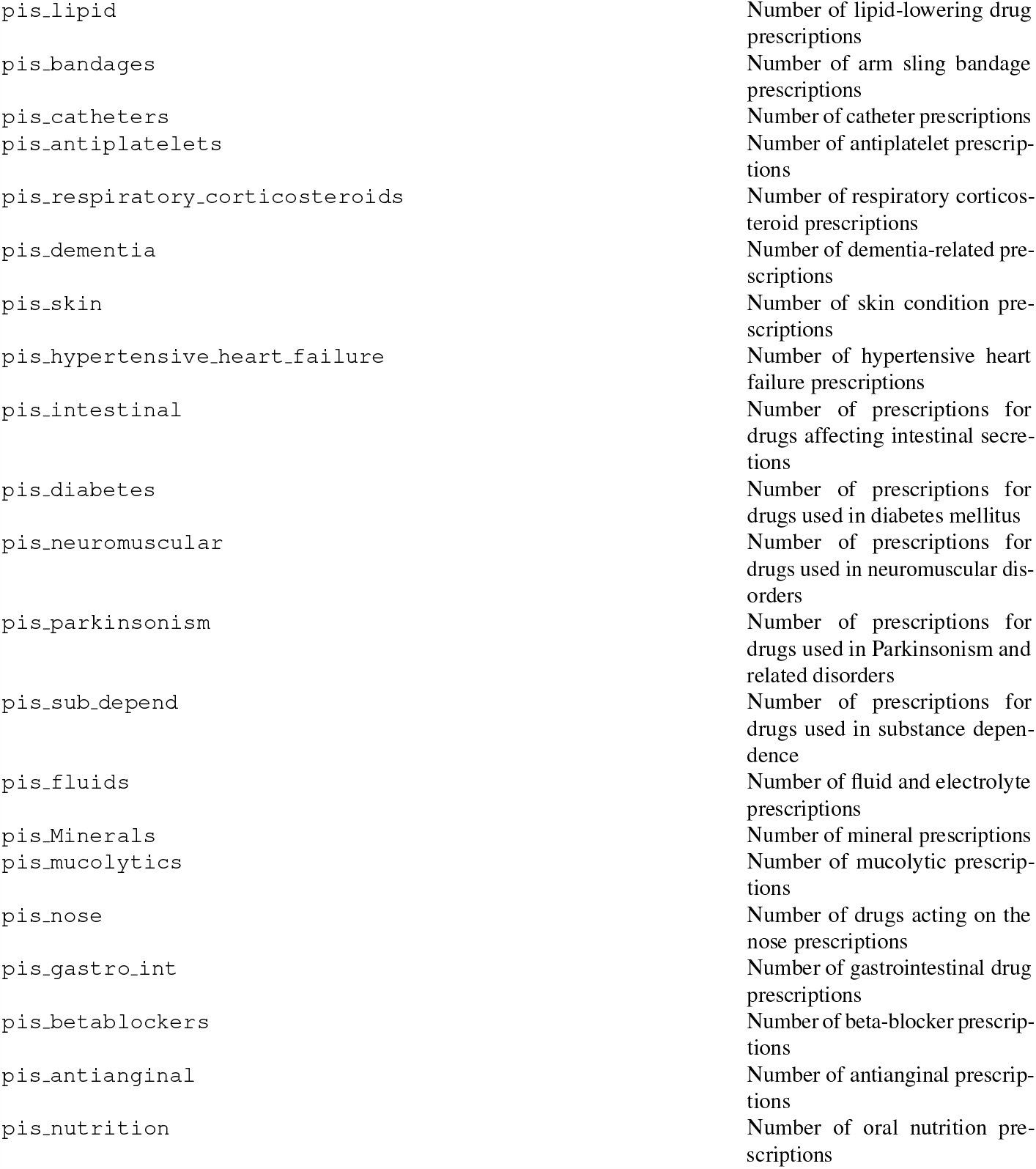
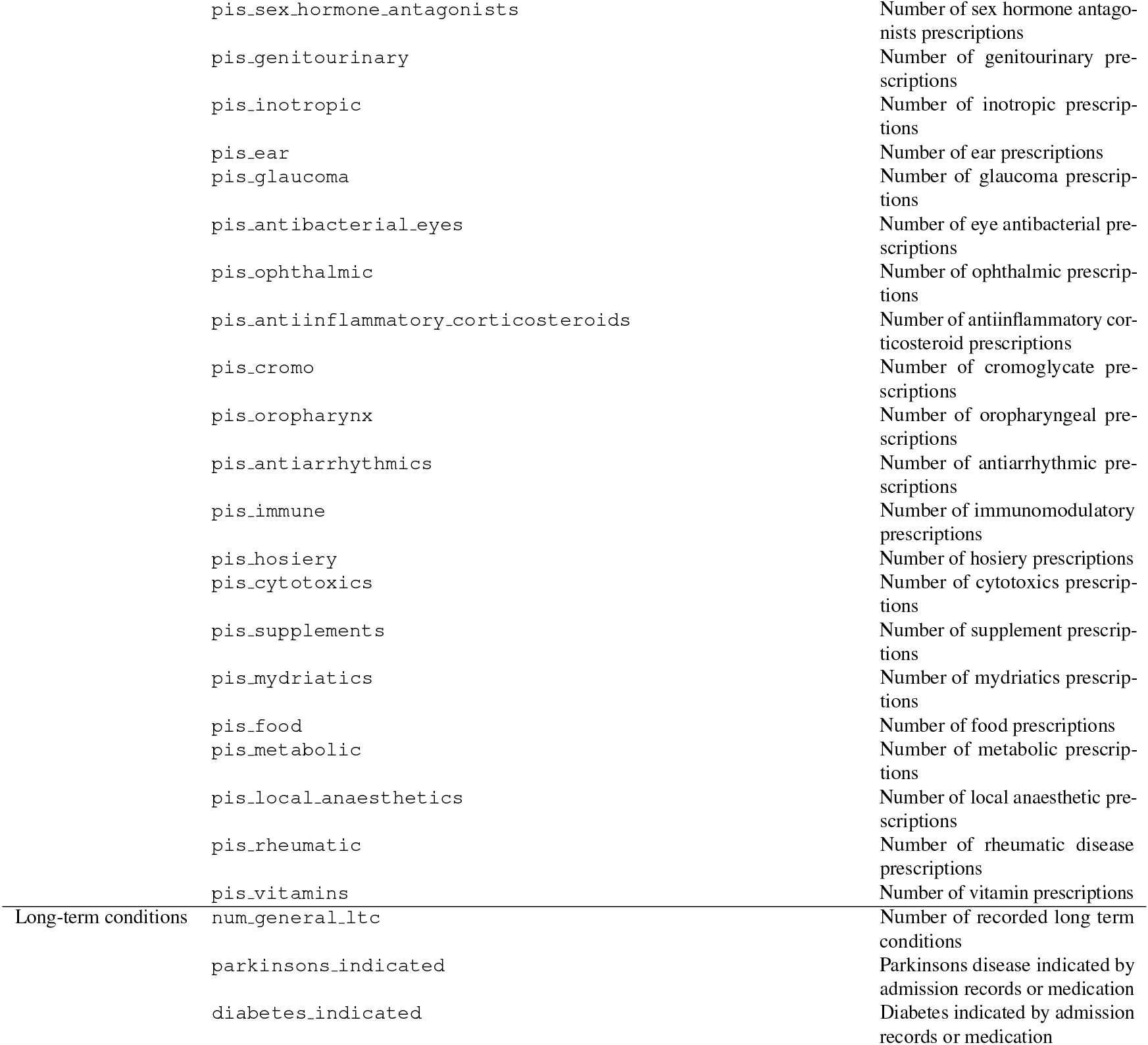
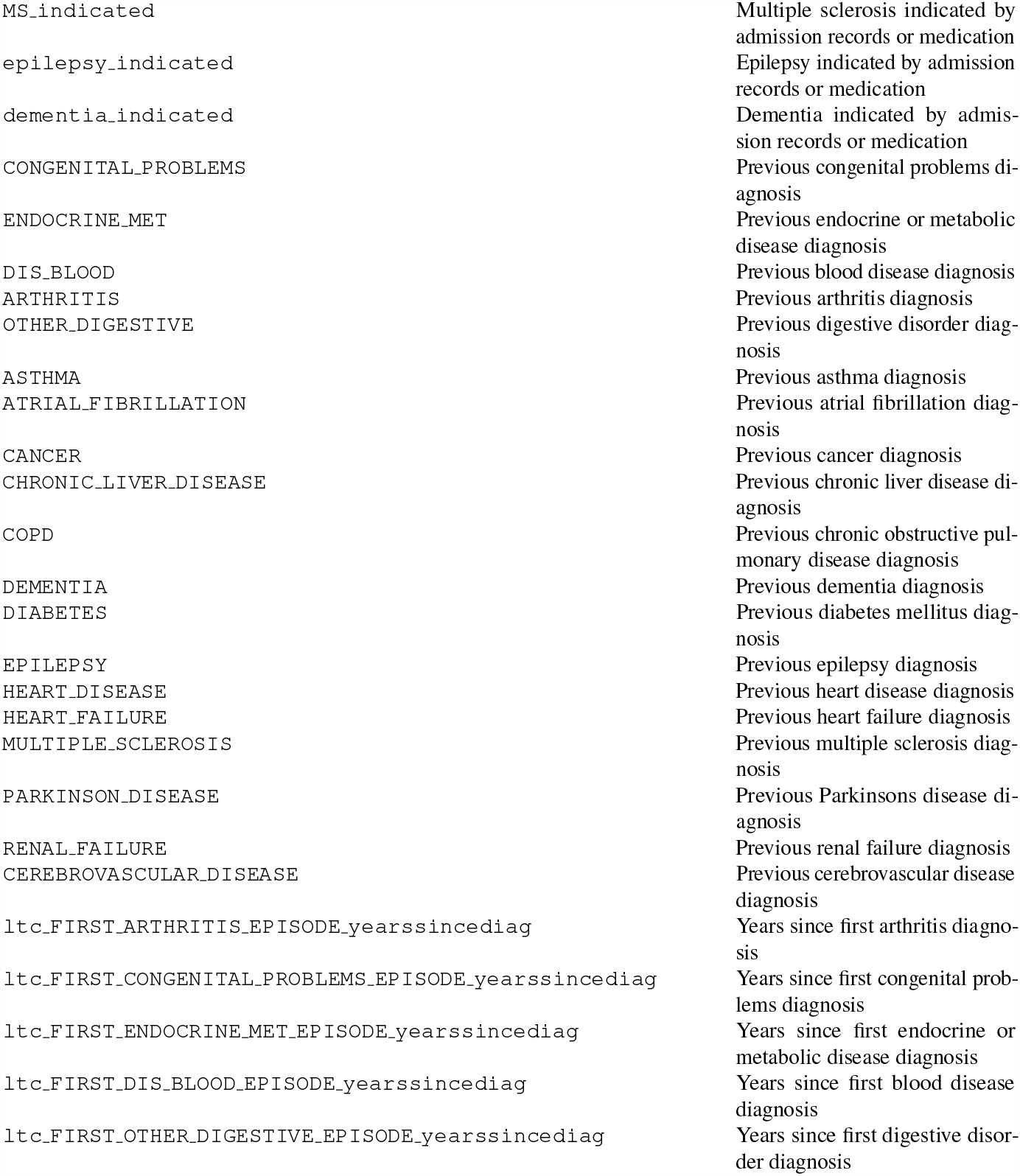
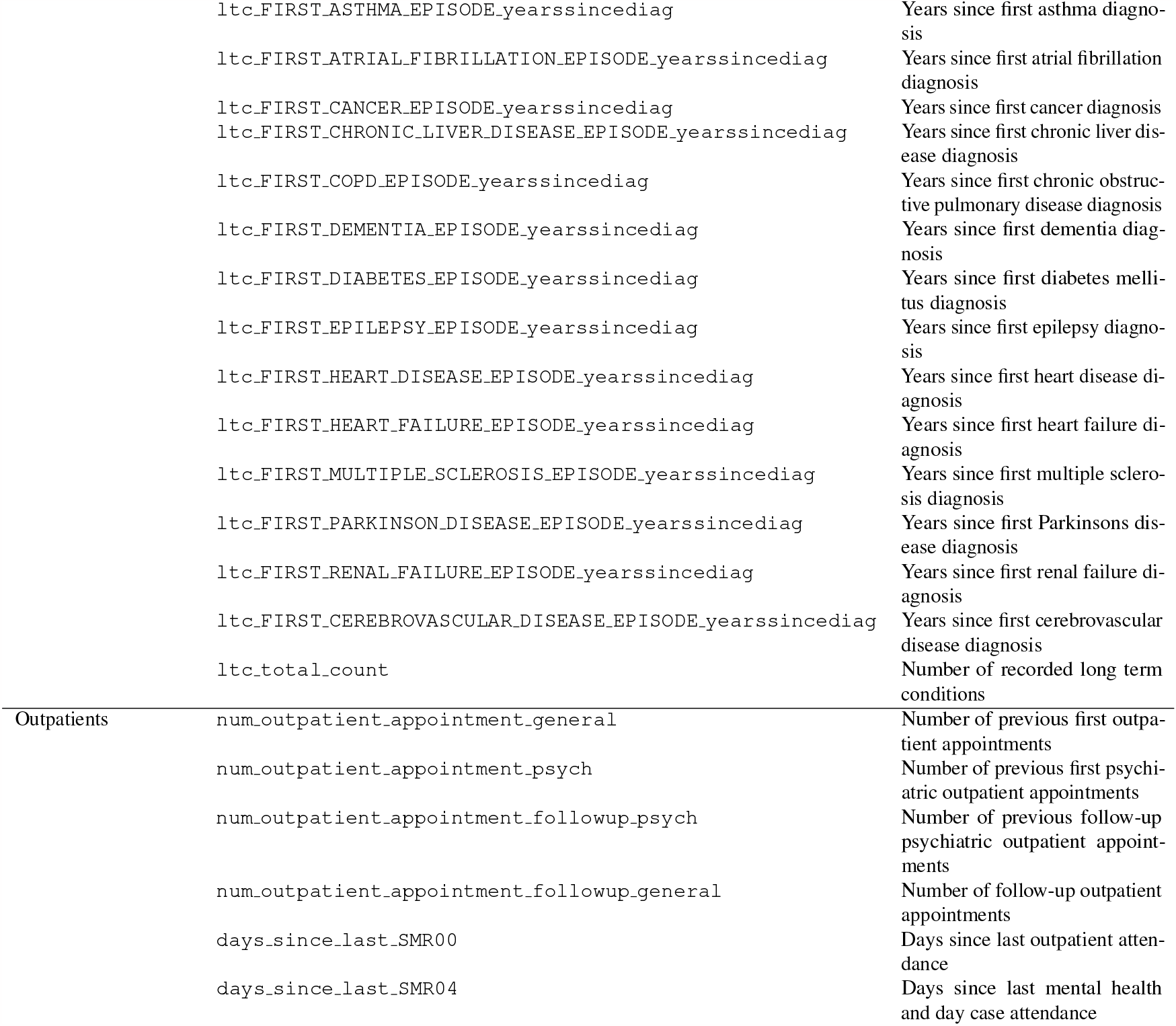
Definition of input features for SPARRAv4. Variable names match the names used in our analysis code.

**Table S4.**
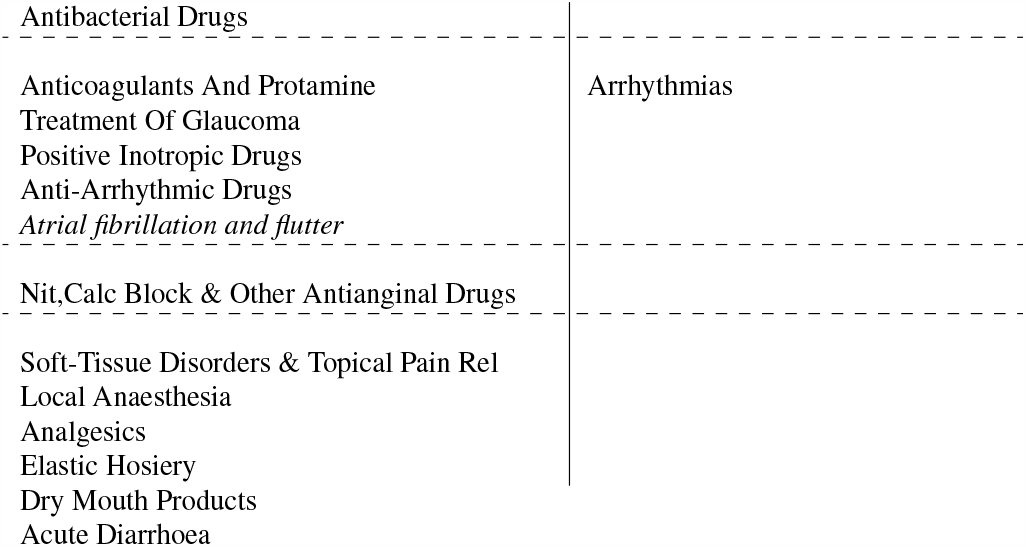
Exploration of the inferred topics. Details of derived topics for topic model used for prediction in F1 (fitted to F2+F3). A topic model assumes that each ‘document’ (individual) in a ‘corpus’ (population) is associated with various ‘topics’ (roughly, illness categories) where each topic corresponds to a distribution over ‘words’ (ICD10 codes and medication types). We would expect that the 30 topics fitted to each fold roughly represent the major clusters of disease types which occur amongst those individuals. This tables shows the ‘words’ with the highest probability of membership in each topic (*>* 1%, where probabilities over all words sum to 100%). In each topic, words are ordered by decreasing probability of topic membership. ICD10 codes are italicised; medication types are not. Topics are ordered by decreasing importance (mean absolute Shapley value). We manually assigned labels to some topics which appear to code for particular disease types.

**Table S5.**
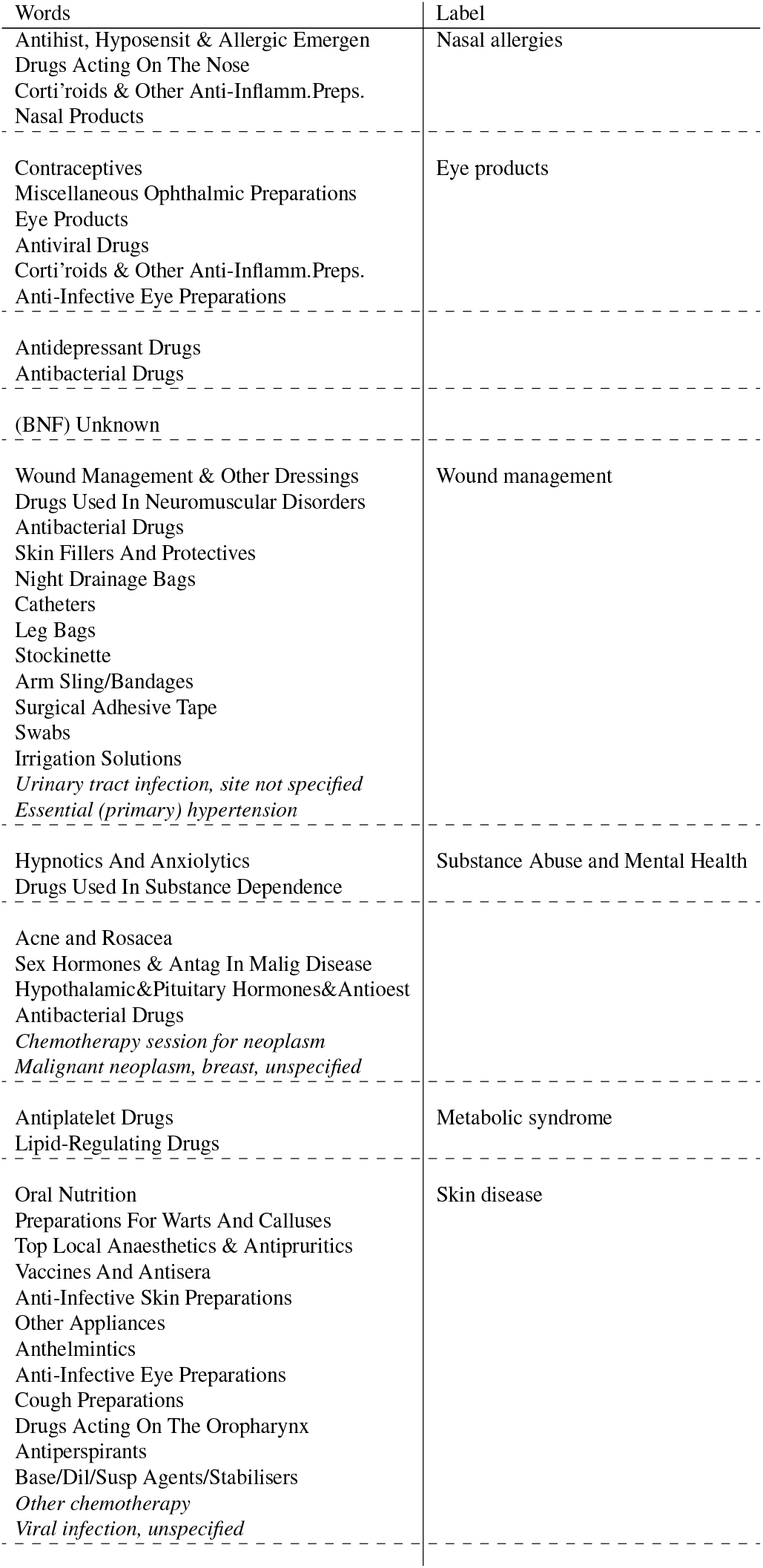
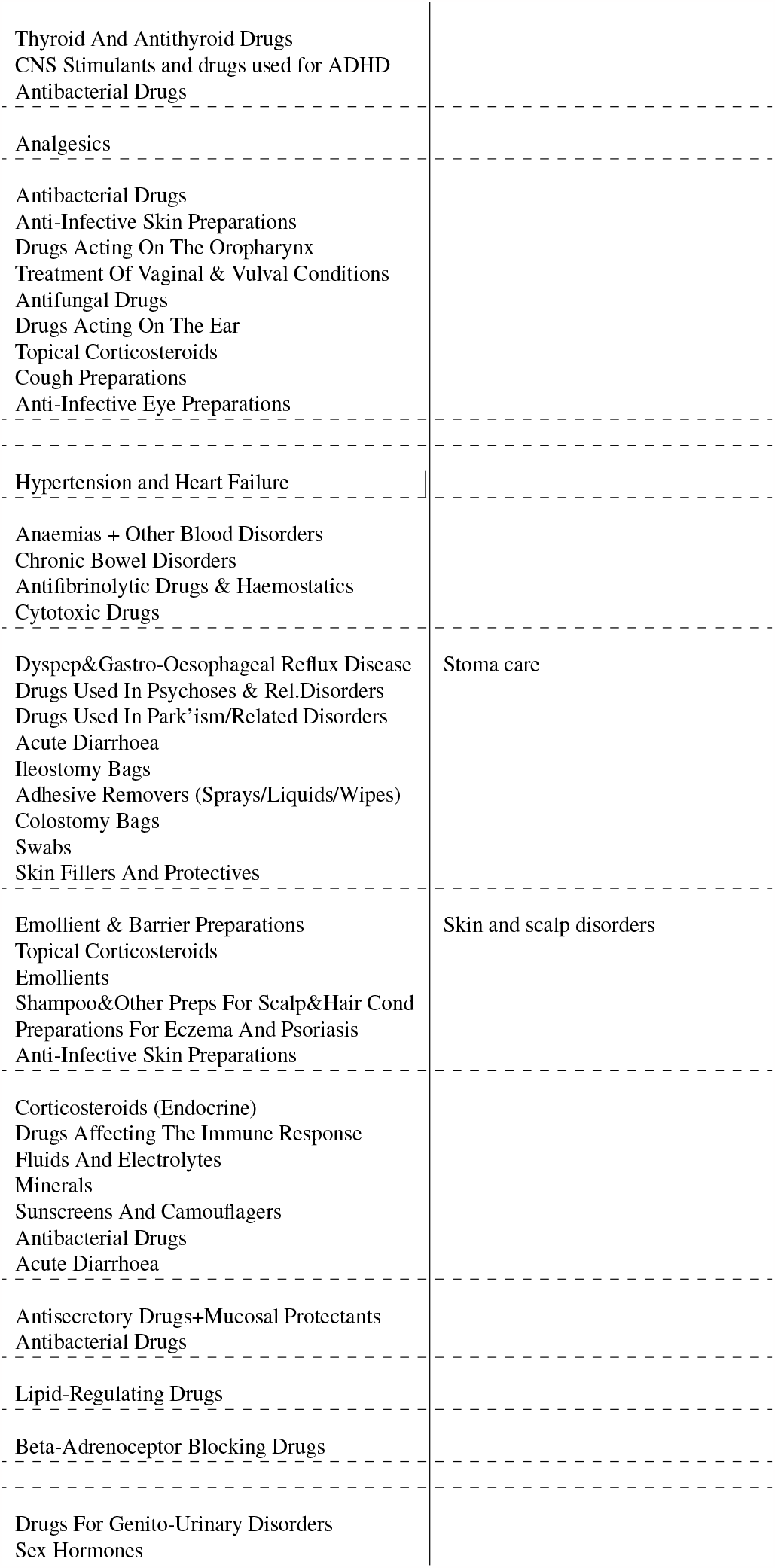
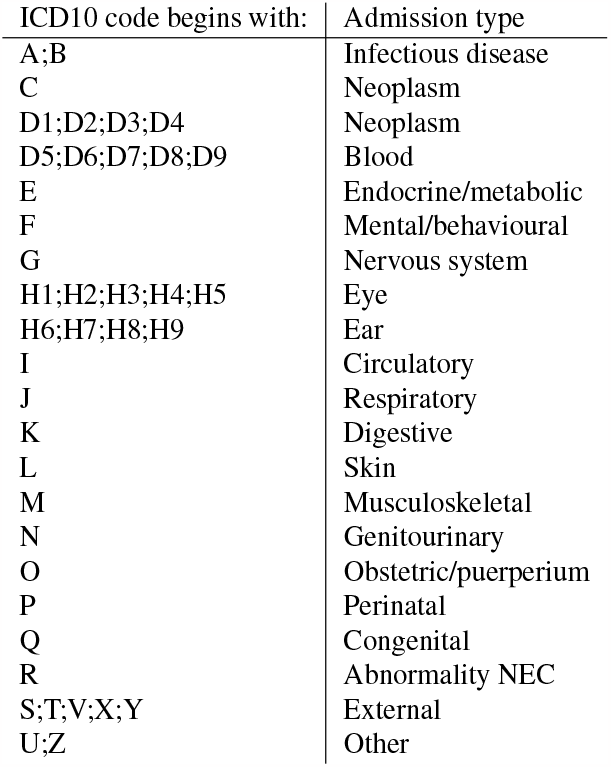
Definition of different admission types.

**Table S6.** Frequency of EA and deaths amongst samples excluded from SPARRAv4. All figures show total numbers. In row names, as for Figure 1B: ‘Died’: Died prior to time cutoff; ‘No SIMD’: missing SIMD; ‘No v3’: missing SPARRAv3 score; ‘Unmatched’: could not match record. For completeness, combinations of exclusions are included even if no individual was excluded with this particular combination.

|  | **Total** | **Admitted only** | **Died** | **Both** |
| --- | --- | --- | --- | --- |
| Excluded | 4,622,512 | 128,241 | 6,139 | 3,166 |
| Included | 12,866,084 | 977,159 | 57,183 | 107,827 |
| Died | 585,322 | 0 | 0 | 0 |
| No v3 | 4,528,514 | 121,857 | 6,036 | 3,011 |
| No SIMD | 199,746 | 12,688 | 151 | 263 |
| Unmatched | 13,524 | 526 | 0 | 0 |
| Died and No v3 | 585,089 | 0 | 0 | 0 |
| Died and No SIMD | 1,258 | 0 | 0 | 0 |
| Died and Unmatched | 0 | 0 | 0 | 0 |
| No v3 and No SIMD | 112,992 | 6,648 | 48 | 110 |
| No v3 and Unmatched | 6,468 | 179 | 0 | 0 |
| No SIMD and Unmatched | 303 | 20 | 0 | 0 |
| Died and No v3 and No SIMD | 1,258 | 0 | 0 | 0 |
| Died and No v3 and Unmatched | 0 | 0 | 0 | 0 |
| Died and No SIMD and Unmatched | 0 | 0 | 0 | 0 |
| No v3 and No SIMD and Unmatched | 258 | 17 | 0 | 0 |
| All | 0 | 0 | 0 | 0 |

**Table S7.** Coefficients of ensemble when fitted separately to SPARRA*v*3 cohorts. Columns YED, U16, FEC, and LTC correspond to subcohorts in SPARRA*v*3; please see Methods.

|  | Fold 1 |  |  |  |
| --- | --- | --- | --- | --- |
|  | YED | U16 | FEC | LTC |
| ANN | 0 | 0 | 0 | 0 |
| Penalised GLM | 0 | 0 | 0 | 0 |
| Naive Bayes | 0 | 0 | 0 | 0 |
| RF, depth: 20 | 0 | 0 | 0.24 | 0.07 |
| RF, depth: 40 | 0.18 | 0.21 | 0.17 | 0.23 |
| SPARRAv3 | 0 | 0 | 0 | 0 |
| XGB depth:3 | 1.86 | 1.57 | 1.24 | 1.69 |
| XGB depth:4 | 0.84 | 1.57 | 0.68 | 0.62 |
| XGB depth: 8 | 3.12 | 3.20 | 2.14 | 2.39 |
|  | Fold 2 |  |  |  |
|  | YED | U16 | FEC | LTC |
| ANN | 0 | 0 | 0 | 0 |
| Penalised GLM | 0 | 0 | 0 | 0 |
| Naive Bayes | 0 | 0 | 0 | 0 |
| RF, depth: 20 | 0 | 0 | 0.09 | 0.01 |
| RF, depth: 40 | 0.22 | 0.20 | 0.24 | 0.16 |
| SPARRAv3 | 0 | 0 | 0 | 0 |
| XGB depth:3 | 1.87 | 1.58 | 1.37 | 1.62 |
| XGB depth:4 | 0.88 | 1.72 | 0.68 | 0.74 |
| XGB depth: 8 | 3.02 | 3.45 | 2.07 | 2.47 |
|  | Fold 3 |  |  |  |
|  | YED | U16 | FEC | LTC |
| ANN | 0 | 0 | 0 | 0 |
| Penalised GLM | 0 | 0 | 0 | 0 |
| Naive Bayes | 0 | 0 | 0 | 0 |
| RF, depth: 20 | 0 | 0 | 0.28 | 0 |
| RF, depth: 40 | 0.16 | 0.11 | 0.15 | 0 |
| SPARRAv3 | 0 | 0 | 0 | 0 |
| XGB depth:3 | 1.73 | 1.16 | 1.31 | 1.60 |
| XGB depth:4 | 1.07 | 1.82 | 0.67 | 0.48 |
| XGB depth: 8 | 3.02 | 3.42 | 2.06 | 2.44 |

## SUPPLEMENTARY FIGURES

**Figure S1.**
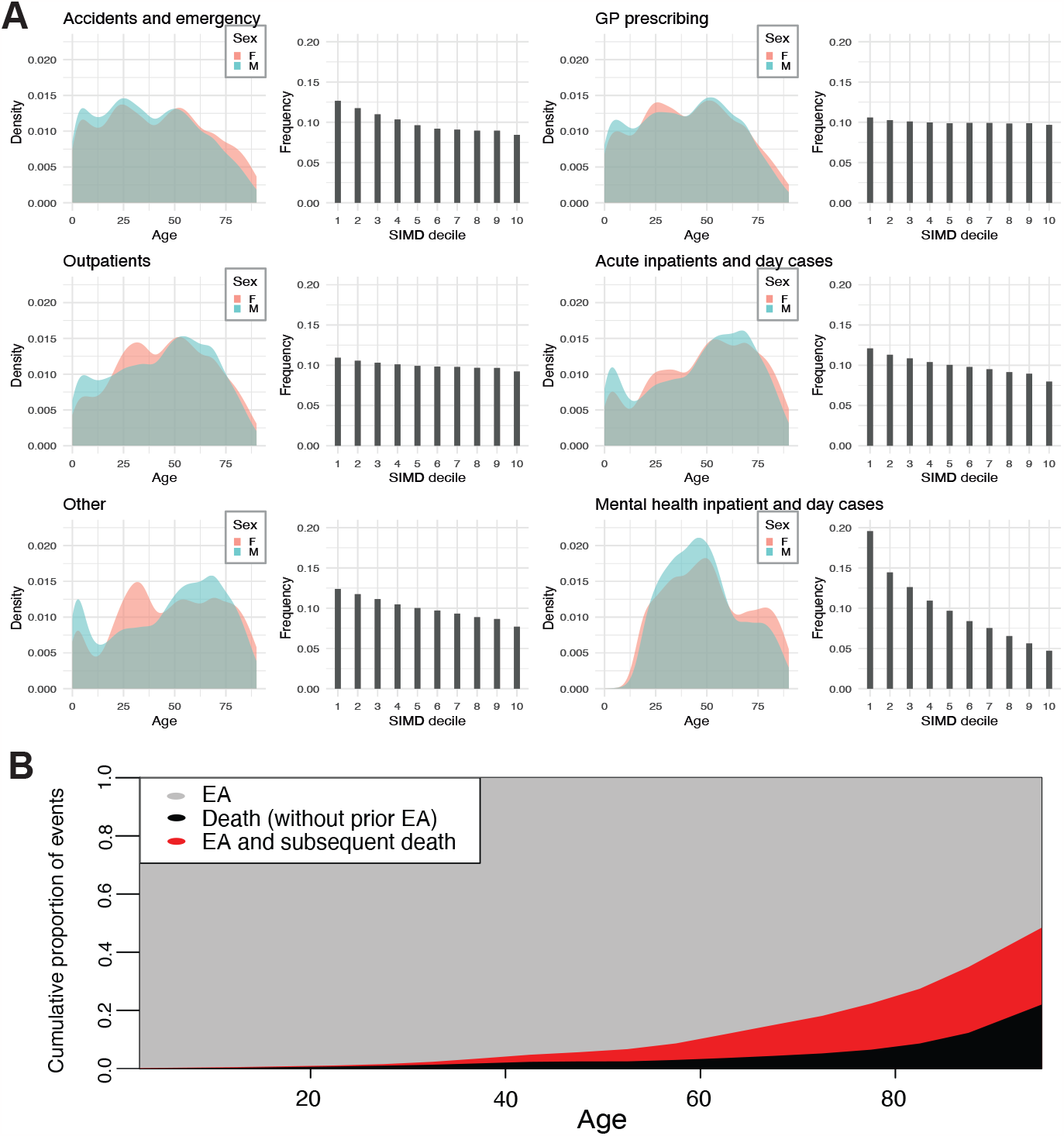
Extended data overview. Distribution of the number of input EHR entries (prior to exclusions) according to age, sex and SIMD deciles (1: most deprived; 10: least deprived) stratified by the input database. All sub-panels are drawn to the same scale. “Other” includes geriatric long stay (SMR01E) and urgent care monitoring (System Watch). (B) Distribution of target events by age stratified by event type.

**Figure S2.**
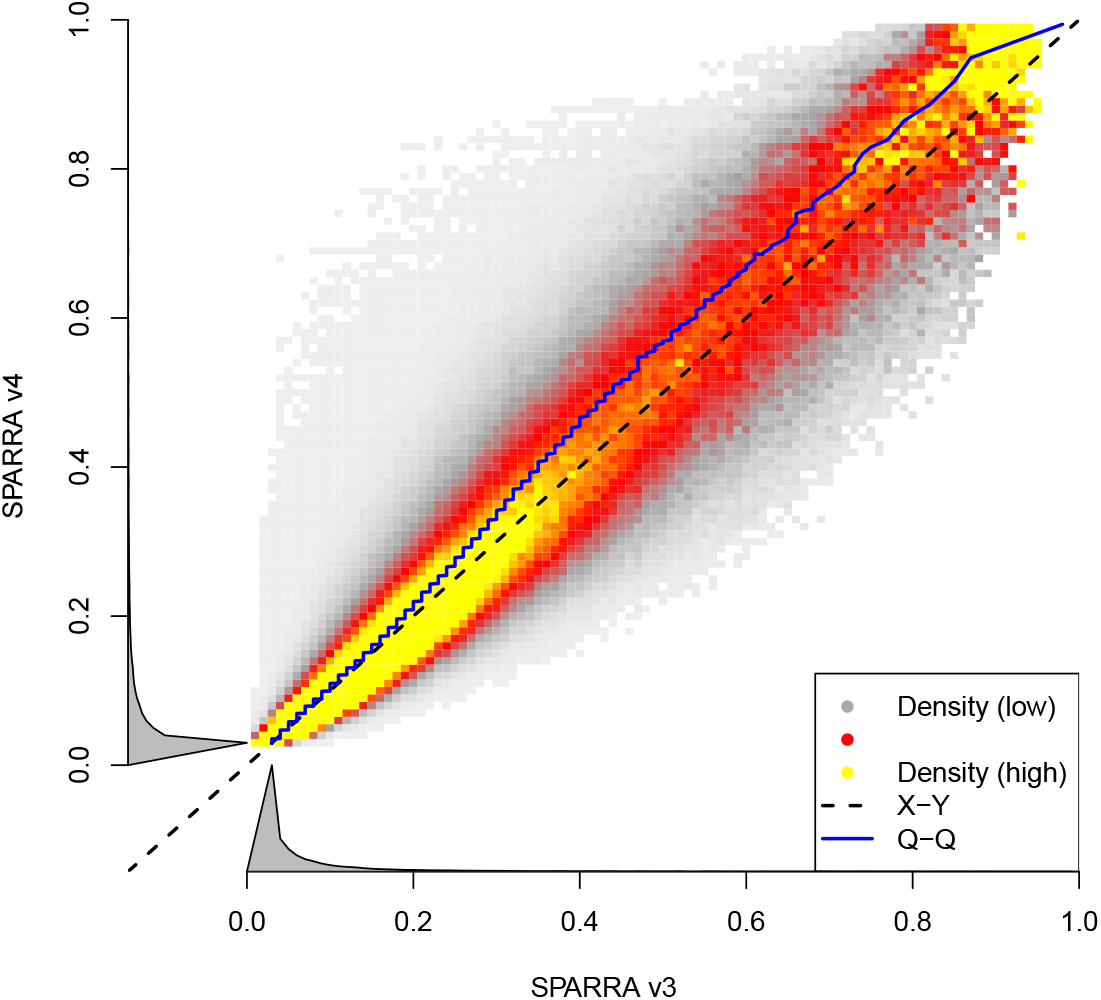
Density plot comparing SPARRAv3 and SPARRAv4 scores. The test datasets used within each CV iteration were combined in order to generate this plot (i.e. all samples are included once). Joint density (low to high: white-grey-red-yellow) of individual SPARRAv3 and SPARRAv4 scores. The density is normalised to uniform marginal on the Y axis, then the X axis; true marginal distributions of risk scores are shown alongside in grey.

**Figure S3.**
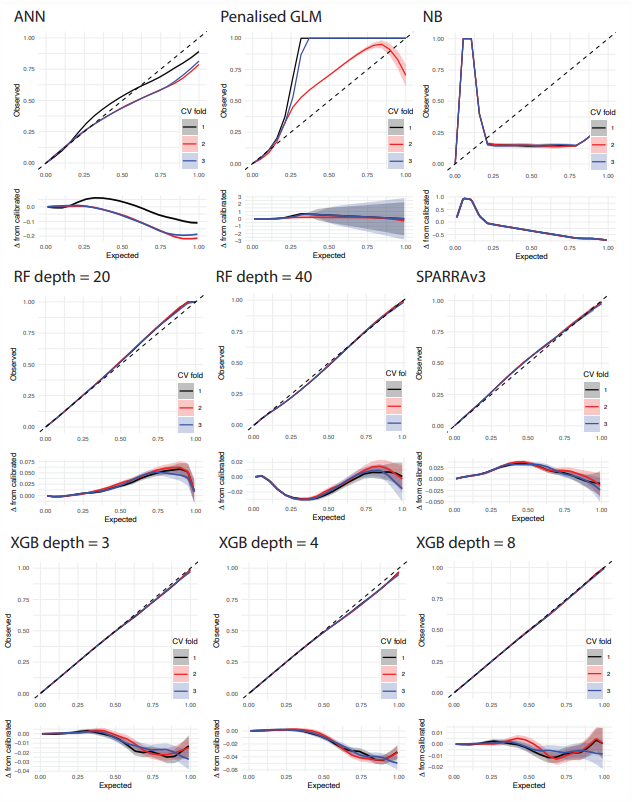
Calibration curves for SPARRAv4 model constituents. Estimates obtained for the test set within each CV iteration are shown in different colours (legend shown in top left panel). Bottom sub-panels show departure from perfect calibration.

**Figure S4.**
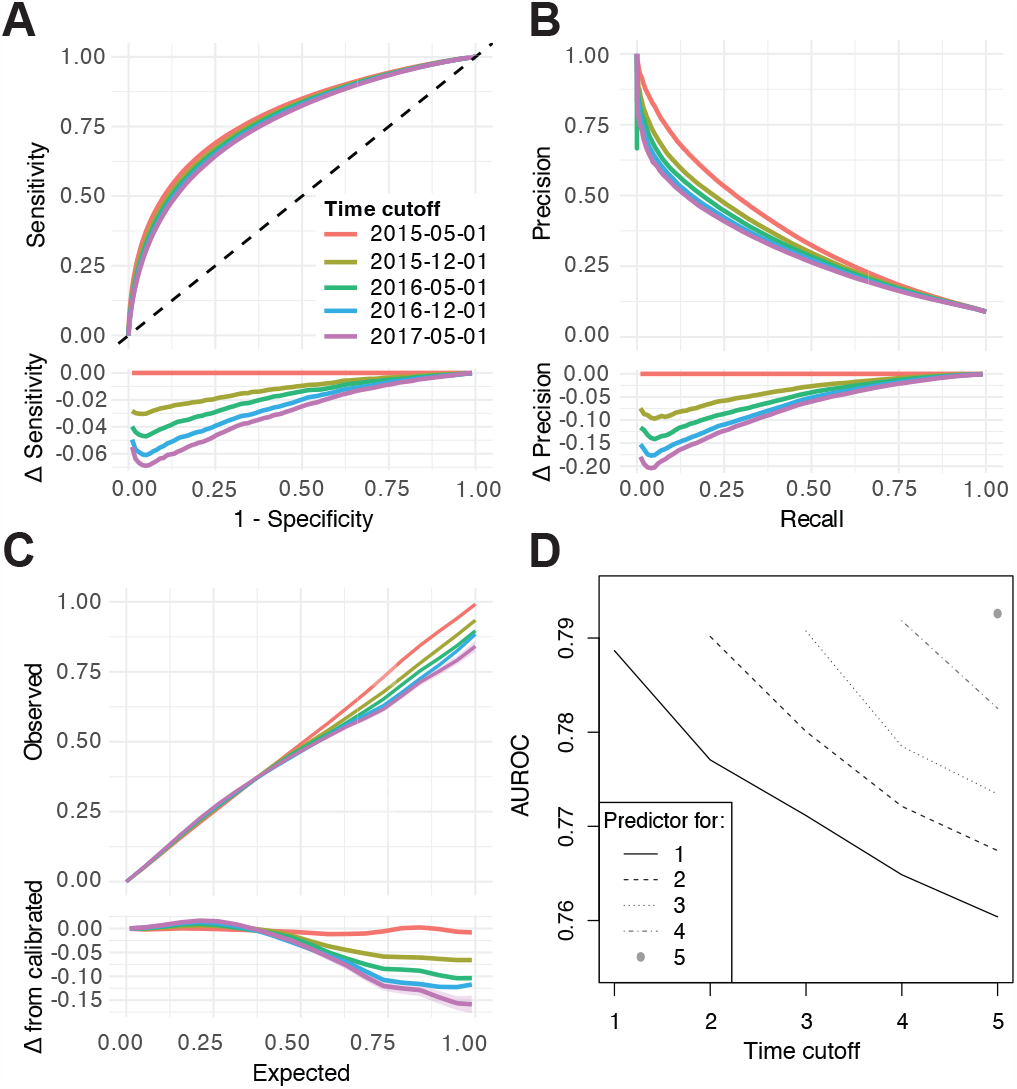
Performance of a static model and static scores used to predict risk at future time cutoffs. (A-C) Performance of static scores evaluated using *M*_0_ at time *t*_0_ for predicting EA at times *t*_1_− *t*_5_. (A) ROC curves. (B) PR curves. (C) Calibration curves. (D) AUROCs for scores calculated at each time cutoff (based on *M*_0_) for prediction in subsequent time cutoffs.

**Figure S5.**
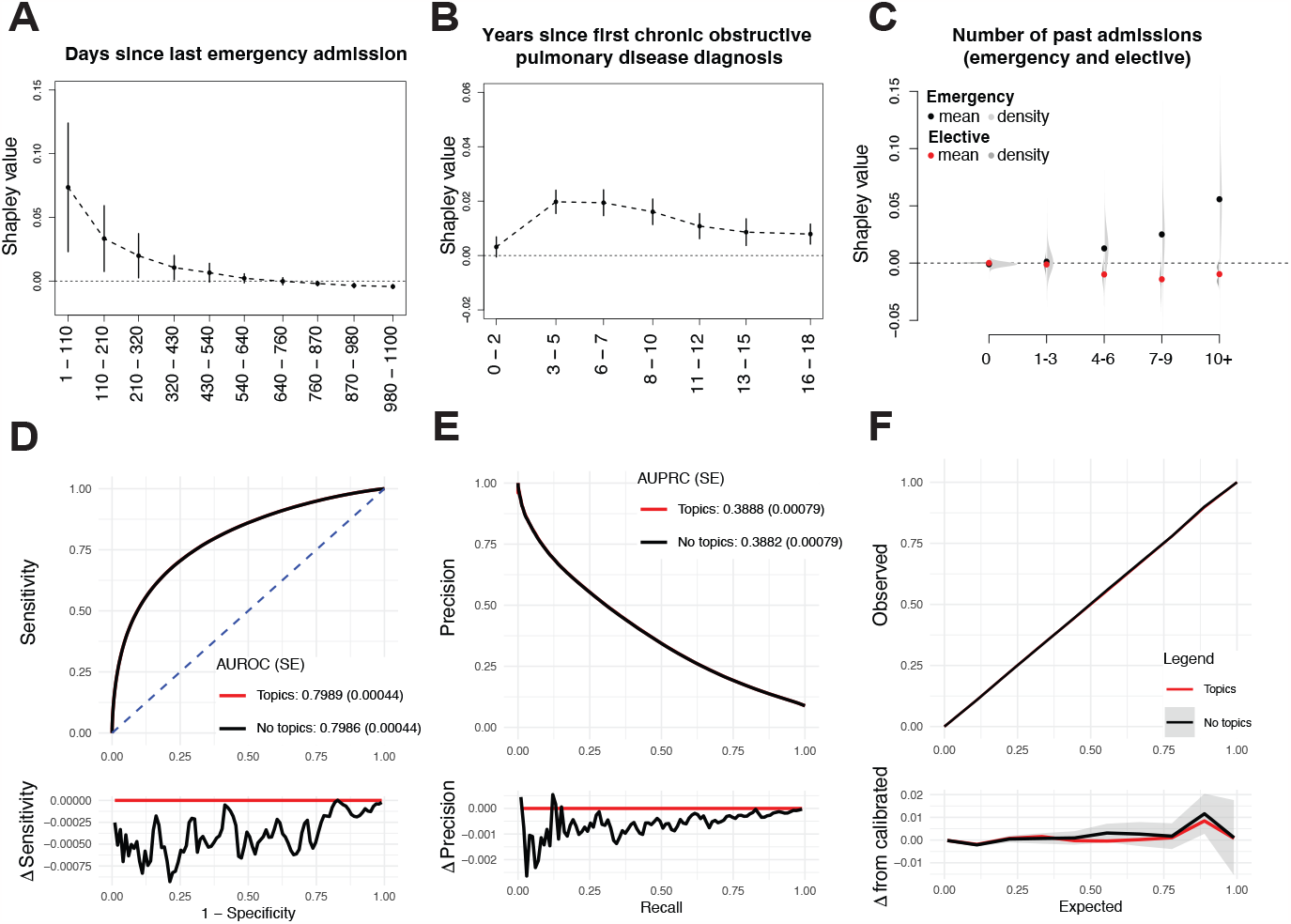
Feature importance. (A-B) Two examples of non-linear feature importance as measured by mean Shapley values (vertical lines show plus/minus one standard deviation). (C) Distribution of Shapley values for the number of previous elective and emergency admissions. (D-E) Comparison of predictive performance with and without topic-model derived features. (D) ROC. (E) PRC. (F) Calibration curve. For (D-E), bottom sub-panels show differences in sensitivity and specificity, respectively. In (F) bottom sub-panel shows difference with respect to perfect calibration.

## SUPPLEMENTARY NOTES

### S1 Analysis of specific topic effects

We searched for individuals for whom the topic model made a substantial difference to their SPARRA score. We considered Shapley values for the 30 topic features across 20,000 uniformly-randomly-chosen individuals in cross-validation fold 1. We searched for topic-derived features for which some of these 20,000 individuals had a Shapley value in excess of 2% for that topic. This is a large Shapley value; for reference, the mean Shapley value from being in the most-deprived decile is only around 1%.

We noted that for one topic feature (topic 21) 87 individuals (about 0.43% of the cohort of 20,000) had a Shapley value in excess of 2%, meaning that the additive contribution to their score from the topic feature was at least 0.02. To gauge the effect of this change, we compared the SPARRA*v*3 and SPARRA*v*4 scores of these individuals (the SPARRA*v*3 score does not use features derived from a topic model). The SPARRA*v*4 scores were on average higher (mean 0.52 for SPARRA*v*4 vs 0.40 for SPARRA*v*3; p-value (t-test) *<* 1 × 10^−4^) and calibration-in-the-large was closer for SPARRA*v*4 (admission frequency 0.56; closer to 0.52 (SPARRA*v*4) than 0.40 (SPARRA*v*3)).

Topic 21 was associated with skin and scalp disorders (see Supplementary Table S4). It is possible that the individiuals for whom this topic feature had a high Shapley value were at elevated risk of EA due to such disorders, but that this was not detectable from the features used in SPARRA*v*3.

In order to avoid data leakage while using our cross-validation scheme, we needed to fit three separate topic models, each fitted to data from two cross-validation folds and used to generate topic features for the remaining fold (see Methods section). For the deployed model, a topic model is refitted to the entire cohort, so the inferred topics are unlikely to contain the exact same cluster of prescriptions and diagnosis. Furthermore, topic-specific contributions may differ from those presented here. However, the analysis above does indicate that, in general, topic features can lead to substantial improvements in score accuracy for some individuals.

### S2 Model re-calibration

We applied a monotonic transformation to optimise the calibration of the scores generated by the ensemble. Given a predicted value 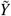 (for ease of notation we do not explicitly include its dependency on the input features *X*) we defined a transformation *m*(·) to optimise calibration, essentially using isotonic regression. The latter was derived using the following procedure.

Fitst, we defined an empirical calibration function for an estimator 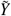 of *Y* |*X* :

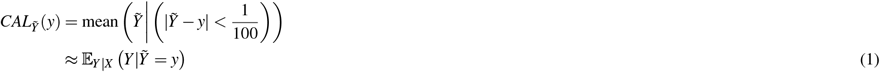

We then found *a, b* such that the mean and mode of 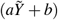 were approximately correctly calibrated; that is, 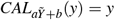 for 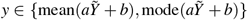, and scaled *a, b* such that 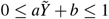. Across an evenly spaced grid *G* of 100 *y*-values we computed the function:

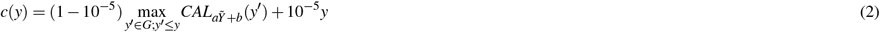

using the cumulative maximum of *CAL*(·) to ensure *c*(·) is non-decreasing, and adding a linear term to ensure *c*(·) is increasing. We extended the domain of *c*(·) to [0, 1] using piecewise linear interpolation, and defined our calibrating transform *m*(·) as the inverse of *c*(·):

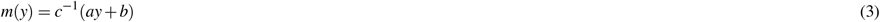

The transformation above was optimised using by further splitting the training set (F1+F2) within our 3-fold cross-validation (CV) procedure (we use F1, F2 and F3 to denote each fold). For each CV-fold, the following steps were performed:

1. Train all constituent models using F1 (except SPARRAv3, for which PHS provided the scores).
2. Each constituent model was then used to generate scores for samples in F2.
3. Given those scores, ensemble weights were inferred via 10-fold CV within F2.
4. Using the previously calculated scores and ensemble weights, the parameters *a* and *b* were chosen to optimise calibration in F2.
5. The optimal ensemble weights and calibration transformation parameters (*a* and *b*) were then used as fixed constants when training the model in the combined F1+F2 dataset.

Note that, due to computational constrains, the topic model was not retrained within the above procedure. Instead, a pre-trained topic model (using F1+F2 as a combined dataset) was used to generate features to be used in step 1.

### S3 Use of subcohort-specific ensemble coefficients

When fitting the SPARRA*v*4 score, for each cross-validation fold, we computed predictions for a range of constituent machine learning models. The final score was defined as a linear combination of the predictions generated by each constituent model a(see Methods section and Table 2). The optimal linear combination was determined by fitting an L-1 penalised generalised linear model with the predictions from the constituent models as input and presence of EA as output.

We considered the possibility that the model could be improved by allowing the coefficients of each model to vary across different subsets of the population. In particular, we assessed the extent to which allowing models to vary across the subcohorts used in the SPARRA*v*3 score (LTC, YED, FEC, and U16; see Methods section) improved the overall performance of the model. We fitted separate penalised regression models (leading to different ensemble weights) in each of the three cross-validation folds and four cohorts (for 12 overall). Subsequently, we assessed how predictive performance changed in the corresponding subcohort in the test set. For instance, we fit one linear model to samples in the YED cohort in folds 2 and 3, and evaluated the performance of this model in samples in the YED cohort in fold 1.

For comparison, we also considered the performance of our original non-cohort-specific ensemble weights, the performance of the best constituent model, and the performance of our original model without topic-model derived features. We evaluated all models using AUROC.

We found that using cohort-specific coefficients in this way improved AUC in the relevant test sets (of 12 comparisons of AUROC, 11 improved; *p <* 0.007 using a Binomial test). However, the magnitude of the change was small: AUC improved in each subcohort by a mean value of only 3.8 × 10^−5^ (where the mean is across cross-validation folds and cohorts). By comparison, use of topic features improves AUC by a mean of 4.1× 10^−4^, around ten times higher, and use of a weighted sum of models rather than just the best-performing constituent model improves AUC by 7.7 × 10^−4^, around twenty times higher.

Given the relatively small advantage of doing this relative to the difficulty of implementation, we opted not to fit separate models in subcohorts in this way. However, this remains an active area of further research.

Table S7 details the coefficients attained in each model. Generally, the same models (XGB and RF) had non-zero coefficients in each case.

### S4 Imputation of lengths of stay when discharge date was missing

Some of our predictors concerned lengths-of-stay; that is, total days spent in hospital in the pre-prediction period (`elective bed days`, `emergency bed days`, and `other bed days`; see Table S3). In general, these were calculated by finding all stays listed for a given individual, subtracting the admission date from the discharge date for each stay, and summing the results across all stays. However, for some hospital stays, no discharge date was present in the source tables. In some cases, this was due to the individual still being in hospital at the time cutoff, but in others was evidently due to the discharge date simply not being recorded; we identified several individuals who were admitted with no discharge date who had evidence of community activity during the time they were supposedly in the hospital. To manage this, we used an imputation procedure for hospital stays in which the discharge date was not recorded. When we see an individual at a time cutoff *t* with admission date *d* and no discharge date, we have options of:

1. Do not count this admission towards the total length of stays; that is, count the stay length as 0 days for that admission. This will under-estimate the total length of stay.
2. Count time *t* − *d* towards the total length of stay. Effectively this imputes the discharge date using the time cutoff. This could lead to incorrect assumptions of very long hospital stays for individuals; indeed, since the pre-prediction period is three years, the mean assumed hospital stay length for such patients would be in excess of eighteen months. This is likely to over-estimate the total length of stay.
3. Count some arbitrary time *t*_0_ towards the total length of stay. Depending on the value of *t*_0_, the total length of stay may be under- or over-estimated.

All of these options coult potentially decrease the usefulness of these variables by artificially inflating (or deflating) the predicted EA risk. As a compromise, we decided to use

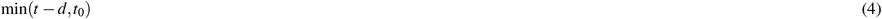

as the length of stay for admissions with a missing discharge date. Effectively, this strategy uses *t*_0_ as a default *minimum length* for stays with missing discharge date.

To choose *t*_0_, we use an empirical Bayes-optimal decision rule. Let *E* be the event that the discharge time for a given admission is not recorded. We model the time *t* − *d* as a (discrete) random variable *X* with a mixture distribution depending on *E*. We want to choose *t*_0_ so that *P*(*E*|*X* = *x*) ≥ 1*/*2 if and only if *x* ≥ *t*_0_. We set

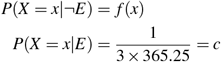

that is; if the discharge time is recorded (in which case the individual is genuinely still in hospital at time *t*), we have some distribution of true lengths of stay, whereas if the discharge time is not recorded, the time *t* − *d* has an equal probability of being anywhere between one day and three years.

Let *P*(*E*) = *q*. Now

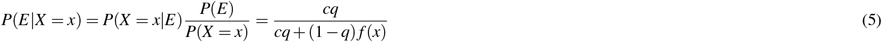

Given estimates of *q* and *f* (·), to find *t*_0_ we may set this expression to 1*/*2 and solve for *x*.

In order to estimate *q* and *f* with 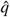 and 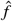, we consider the population P of admissions (not individuals) where the admission date is between May 2013 and May 2014. We then estimate

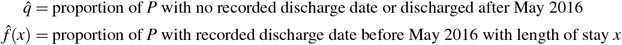

We use this population of admissions so as to avoid data leakage, since these are prior to the earliest time cutoff (May 2016) used in fitting the model. This is also our rationale for treating individuals who were discharged post-May 2016 the same as having no recorded discharge date: we cannot use this information without data leakage. However, we note that the number of individuals with genuine *>* 2 year hospital stays is very small.

Following this procedure, estimated values of *t*_0_ are 26, 19 and 6 for `emergency bed days`, `elective bed days` and `other bed days`, respectively.

### S5 Assessment of calibration

We use an estimator for calibration broadly based on the Nadaraya-Watson kernel estimator [Nadaraya, 1964, Watson, 1964]. We re-derive several properties (consistency, bias) to highlight their interpretation in our context.

We assume in general that, for IID predictor/outcome pairs (*X*_*i*_,*Y*_*i*_) ∼ (*X,Y*), *i* ∈ 1..*n*, and an optimal predictor function *p*_*opt*_, we have

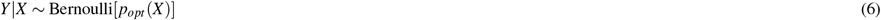

noting that this implies

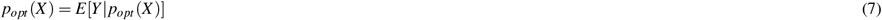

We want to estimate *p*_*opt*_ (*X*).

Since we only observe *Y* = 1 or *Y* = 0, we must estimate *E*[*Y*| *p*(*X*) = *z*] as some kind of average of *Y* about observed values *p*(*X*) close to *z*. A routine way to do this is to use ‘reliability diagrams’ [Bröcker and Smith, 2007] in which we bin values of *p*(*X*) and estimate *E*(*Y* |*p*(*X*)) in each bin.

Since for small bin sizes there may be few or no values of *p*(*X*) in some bins, we use a kernel estimate *ĉ*_*p*_(*z*) of *c*_*p*_(*z*) = *E*[*Y* |*p*(*X*) = *z*]:

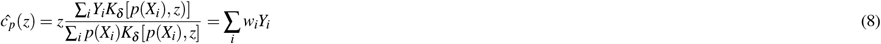

where *K*_*δ*_ : (0, 1)^2^ → ℝ^+^ is some distance-measuring kernel with width *δ*, and

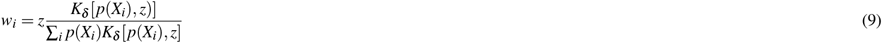

We avoid the simpler estimate given by the *K*_*δ*_ -weighted mean of *Y*_*i*_s:

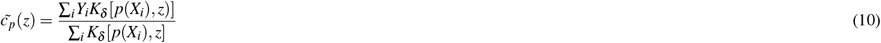

for reasons shown below. We note the following:

#### Proposition 1.

*If p*(*X*) *has Lesbegue-integrable positive density on* (0, 1), *K*(*z, x*) *and c*_*p*_(*x*) *are Lesbegue-integrable functions of x for fixed z >* 0, *and the kernel ‘narrows with δ’ so*

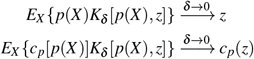

*then ĉ*(*z*) *becomes a consistent estimator of c*(*z*) *as δ* → 0

*Proof*. From Slutsky’s lemma, the law of total expectation and the strong law of large numbers

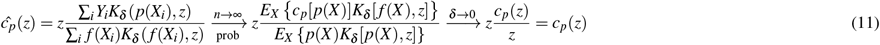

We note that *ĉ*_*p*_(*Z*) is not generally consistent if *δ >* 0. However, the inconsistency is not severe: we note □

#### Proposition 2.

*If, in addition to the above, K*_*δ*_ (*x, z*) = *K*_*δ*_ (*x* − *z*) *is a symmetric density with second moment δ and negligible moments of higher order, and the densities of p*(*X*) *and c*_*p*_(*X*) *are twice differentiable at z, then ĉ*_*p*_(*z*) → *c*_*p*_(*z*) + *O*(*δ* ^2^)

*Proof*. We have

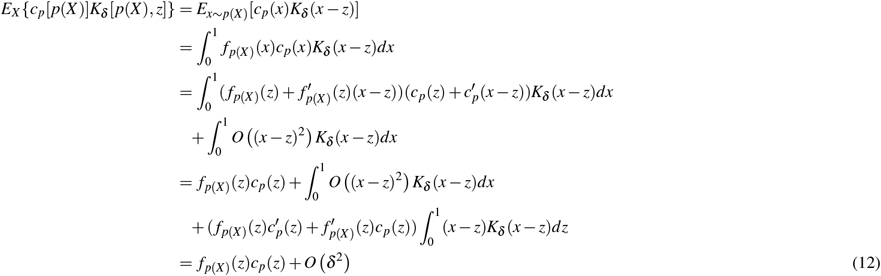

noting the symmetry of *K*_*δ*_. If we replace *c*_*p*_[*p*(*X*)] with *p*(*X*), the expectation is *z f*_*p*(*X*)_(*z*) + *O*(*δ* ^2^), and the result follows from the first part of 11. □

#### Remark 1.

*In the ideal case where c*_*p*_(*z*) = *z (that is, our model is perfectly calibrated) estimator 9 is consistent even when δ >* 0, *whereas the apparently simpler asymptotically consistent (as δ* → 0*) estimator of a weighted sum of Y*_*i*_*’s:*

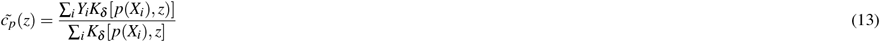

*is not*.

Finally, we note the following:

#### Proposition 3.

*Under the assumptions above, with fixed X*_*i*_, *the bias of* 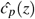 *is*

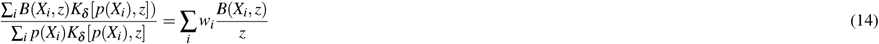

*where B*(*X*_*i*_, *z*) = *p*(*X*_*i*_)*c*_*p*_(*z*) − *zc*_*p*_(*p*(*X*_*i*_)).

*Proof*. With fixed *X*_*i*_

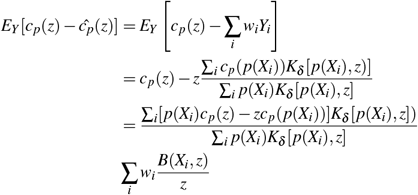

as required. □

#### Remark 2.

*This enables straightforward evaluation of bounds on bias given bounds on the form of c*_*p*_. *The estimator ĉ*_*p*_ *is unbiased if c*_*p*_(*x*) = *kx for some k, since B*(*X*_*i*_, *z*) = 0.

#### Remark 3.

*An alternative way to draw a kernelised calibration curve is to simply plot a parametric curve*

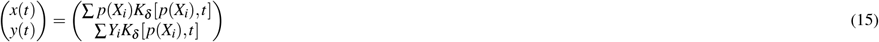

*which, for each t, is an only-slightly biased estimate of some point z, c*_*p*_(*z*). *If a rectangular kernel is used, this is equivalent to binning values of p*(*X*_*i*_) *[Bröcker and Smith, 2007]. However, this method does not generally give a curve across the entire range of p*(*X*_*i*_).

It is straightforward to estimate

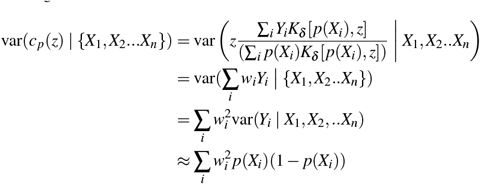

where the approximation is exact if *c*_*p*_(*z*) = *z*. Together with an estimate of maximum absolute bias *b*_*z*_ at *z*, this enables estimates of conservative confidence intervals on *ĉ*_*p*_(*z*) at level 1 − *α* :

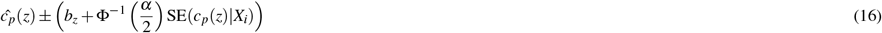

In all plots in this paper, we bounded bias under the assumption that there existed *k* such that |*c*_*p*_(*z*) −*kz* |*< z*^2^*/*10.

The calibration estimator derived here is demonstrated in an R script `sparra calibration.R` available with the attached R code for this manuscript.

## References

Denis Agniel, Isaac S Kohane, and Griffin M Weber. Biases in electronic health record data due to processes within the healthcare system: retrospective observational study. Bmj, 361, 2018.

N Bajaj, S Jauhar, and J Taylor. Scottish patients at risk of readmission and admission-mental health (SPARRA MH) case study of users and non-users of a national information source. Health Syst Policy Res, 3:3, 2016.

Stephen Bates, Trevor Hastie, and Robert Tibshirani. Cross-validation: what does it estimate and how well does it do it? Journal of the American Statistical Association, pages 1–12, 2023.

John Billings, Jennifer Dixon, Tod Mijanovich, and David Wennberg. Case finding for patients at risk of readmission to hospital: development of algorithm to identify high risk patients. BMJ, 333(7563):327, 2006.

David M Blei, Andrew Y Ng, and Michael I Jordan. Latent Dirichlet allocation. Journal of machine Learning research, 3(Jan):993–1022, 2003.

Ian Blunt. Focus on preventable admissions. London: Nuffield Trust, 2013.

Alex Bottle, Paul Aylin, and Azeem Majeed. Identifying patients at high risk of emergency hospital admissions: a logistic regression analysis. Journal of the Royal Society of Medicine, 99(8):406–414, 2006.

Anne Canny, Frances Robertson, Peter Knight, Adam Redpath, and Miles D Witham. An evaluation of the psychometric properties of the indicator of relative need (IoRN) instrument. BMC geriatrics, 16(1):1–10, 2016.

Tianqi Chen, Tong He, Michael Benesty, Vadim Khotilovich, Yuan Tang, Hyunsu Cho, Kailong Chen, Rory Mitchell, Ignacio Cano, Tianyi Zhou, Mu Li, Junyuan Xie, Min Lin, Yifeng Geng, and Yutian Li. xgboost: Extreme Gradient Boosting, 2019. URL https://CRAN.R-project.org/package=xgboost. R package version 0.90.0.2.

Joanna Coast, Abby Inglis, and Stephen Frankel. Alternatives to hospital care: what are they and who should decide? BMJ, 312(7024):162–166, 1996.

Gary S Collins, Johannes B Reitsma, Douglas G Altman, and Karel GM Moons. Transparent reporting of a multivariable prediction model for individual prognosis or diagnosis (tripod): the tripod statement. Journal of British Surgery, 102(3):148–158, 2015.

Elizabeth R DeLong, David M DeLong, and Daniel L Clarke-Pearson. Comparing the areas under two or more correlated receiver operating characteristic curves: a nonparametric approach. Biometrics, pages 837–845, 1988.

David A Ellis, Ross McQueenie, Alex McConnachie, Philip Wilson, and Andrea E Williamson. Demographic and practice factors predicting repeated non-attendance in primary care: a national retrospective cohort analysis. The Lancet Public Health, 2(12): e551–e559, 2017.

George Forman and Martin Scholz. Apples-to-apples in cross-validation studies: pitfalls in classifier performance measurement. Acm Sigkdd Explorations Newsletter, 12(1):49–57, 2010.

Health and Social Care Information Programme. A report on the development of SPARRA version 3 (developing risk prediction to support preventative and anticipatory care in Scotland), 2011. https://www.isdscotland.org/Health-Topics/Health-and-Social-Community-Care/SPARRA/2012-02-09-SPARRA-Version-3.pdf, Accessed: 6-3-2020.

Gill Highet, Debbie Crawford, Scott A Murray, and Kirsty Boyd. Development and evaluation of the supportive and palliative care indicators tool (SPICT): a mixed-methods study. BMJ supportive & palliative care, 4(3):285–290, 2014.

Julia Hippisley-Cox and Carol Coupland. Predicting risk of emergency admission to hospital using primary care data: derivation and validation of QAdmissions score. BMJ open, 3 (8):e003482, 2013.

ISD Scotland Data Dictionary. CHI - Community Health Index, 2023. https://www.ndc.scot.nhs.uk/Dictionary-A-Z/Definitions/index.asp?ID=128, Accessed: 17-3-2023.

Emily Jefferson, James Liley, Maeve Malone, Smarti Reel, Alba Crespi-Boixader, Xaroula Kerasidou, Francesco Tava, Andrew McCarthy, Richard Preen, Alberto Blanco-Justicia, et al. GRAIMATTER green paper: Recommendations for disclosure control of trained machine learning (ML) models from trusted research environments (TREs). arXiv preprint arXiv:2211.01656, 2022.

Ron Kremer, Syed Mohib Raza, Fabiola Eto, John Casement, Christian Atallah, Sarah Finer, Dennis Lendrem, Michael Barnes, Nick J Reynolds, and Paolo Missier. Tracking trajectories of multiple long-term conditions using dynamic patient-cluster associations. In 2022 IEEE International Conference on Big Data (Big Data), pages 4390–4399. IEEE, 2022.

Attakrit Leckcivilize, Paul McNamee, Christopher Cooper, and Robby Steel. Impact of an anticipatory care planning intervention on unscheduled acute hospital care using difference-in-difference analysis. BMJ health & care informatics, 28(1), 2021.

Erin LeDell, Navdeep Gill, Spencer Aiello, Anqi Fu, Arno Candel, Cliff Click, Tom Kraljevic, Tomas Nykodym, Patrick Aboyoun, Michal Kurka, and Michal Malohlava. h2o: R Interface for ‘H2O’, 2019. URL https://CRAN.R-project.org/package=h2o. R package version 3.26.0.2.

Matthew C Lenert, Michael E Matheny, and Colin G Walsh. Prognostic models will be victims of their own success, unless… . Journal of the American Medical Informatics Association, 26(12):1645–1650, 2019.

James Liley, Samuel R Emerson, Bilal A Mateen, Catalina A Vallejos, Louis JM Aslett, and Sebastian J Vollmer. Model updating after interventions paradoxically introduces bias. AISTATS proceedings, 2021.

Scott M Lundberg and Su-In Lee. A unified approach to interpreting model predictions. In Advances in neural information processing systems, pages 4765–4774, 2017.

David Lyon, Gillian A Lancaster, Steve Taylor, Chris Dowrick, and Hannah Chellaswamy. Predicting the likelihood of emergency admission to hospital of older people: development and validation of the emergency admission risk likelihood index (EARLI). Family practice, 24(2):158–167, 2007.

S Manoukian, S Stewart, N Graves, H Mason, C Robertson, S Kennedy, J Pan, L Haahr, SJ Dancer, B Cook, et al. Evaluating the post-discharge cost of healthcare-associated infection in NHS Scotland. Journal of Hospital Infection, 114:51–58, 2021.

Matthew BA McDermott, Shirly Wang, Nikki Marinsek, Rajesh Ranganath, Luca Foschini, and Marzyeh Ghassemi. Reproducibility in machine learning for health research: Still a ways to go. Science Translational Medicine, 13(586):eabb1655, 2021.

Marian S McDonagh, David H Smith, and Maria Goddard. Measuring appropriate use of acute beds: a systematic review of methods and results. Health policy, 53(3):157–184, 2000.

NICE guidelines. Asthma: diagnosis, monitoring and chronic asthma management. National Institute of Health and Care Excellence, November 2017.

Office for National Statistics, National Records of Scotland, and Northern Ireland Statistics and Research Agency. 2011 census aggregate data. UK data service (edition: June 2011), 2011.

Juan Perdomo, Tijana Zrnic, Celestine Mendler-Duünner, and Moritz Hardt. Performative prediction. In International Conference on Machine Learning, pages 7599–7609. PMLR, 2020a.

Juan Perdomo, Tijana Zrnic, Celestine Mendler-Duünner, and Moritz Hardt. Performative prediction. In International Conference on Machine Learning, pages 7599–7609. PMLR, 2020b.

Anne B Prasad. British National Formulary. Psychiatric Bulletin, 18(5):304–304, 1994.

Public Health Scotland. eDRIS Products and Services, Public Health Scotland, 2020. URL https://www.isdscotland.org/Products-and-Services/eDRIS/.

Public Health Scotland. Acute hospital activity and NHS beds information for Scotland, 2022. URL https://publichealthscotland.scot/media/15288/2022-09-27-annual-acuteactivity-report.pdf.

Fatemeh Rahimian, Gholamreza Salimi-Khorshidi, Amir H Payberah, Jenny Tran, Roberto Ayala Solares, Francesca Raimondi, Milad Nazarzadeh, Dexter Canoy, and Kazem Rahimi. Predicting the risk of emergency admission with machine learning: Development and validation using linked electronic health records. PLoS medicine, 15(11): e1002695, 2018.

Rafael D Romo, Theresa A Allison, Alexander K Smith, and Margaret I Wallhagen. Sense of control in end-of-life decision-making. Journal of the American Geriatrics Society, 65 (3):e70–e75, 2017.

Rural Access Action Team. The national framework for service change in NHS Scotland. Scottish Executive, Edinburgh, 2005.

Colin Sanderson and Jennifer Dixon. Conditions for which onset or hospital admission is potentially preventable by timely and effective ambulatory care. Journal of health services research & policy, 5(4):222–230, 2000.

Scottish Government. Scottish index of multiple deprivation, 2016.

Matthew Sperrin, David Jenkins, Glen P Martin, and Niels Peek. Explicit causal reasoning is needed to prevent prognostic models being victims of their own success. Journal of the American Medical Informatics Association, 26(12):1675–1676, 2019.

Adarsh Subbaswamy and Suchi Saria. From development to deployment: dataset shift, causality, and shift-stable models in health ai. Biostatistics, 21(2):345–352, 2020.

Mark J Van der Laan, Eric C Polley, and Alan E Hubbard. Super learner. Statistical applications in genetics and molecular biology, 6(1), 2007.

Robert A Verheij, Vasa Curcin, Brendan C Delaney, and Mark M McGilchrist. Possible sources of bias in primary care electronic health record data use and reuse. Journal of medical Internet research, 20(5):e185, 2018.

Emma Wallace, Ellen Stuart, Niall Vaughan, Kathleen Bennett, Tom Fahey, and Susan M Smith. Risk prediction models to predict emergency hospital admission in community-dwelling adults: a systematic review. Medical care, 52(8):751, 2014.

Emma Wallace, Susan M Smith, Tom Fahey, and Martin Roland. Reducing emergency admissions through community based interventions. BMJ, 352, 2016.

World Health Organization. International statistical classification of diseases and related health problems, volume 1. World Health Organization, 2004.

## REFERENCES

Jochen Bröcker and Leonard A Smith. Increasing the reliability of reliability diagrams. Weather and forecasting, 22(3):651–661, 2007.

Karel GM Moons, Douglas G Altman, Johannes B Reitsma, John PA Ioannidis, Petra Macaskill, Ewout W Steyerberg, Andrew J Vickers, David F Ransohoff, and Gary S Collins. Transparent reporting of a multivariable prediction model for individual prognosis or diagnosis (TRIPOD): explanation and elaboration. Annals of internal medicine, 162(1):W1–W73, 2015.

Elizbar A Nadaraya. On estimating regression. Theory of Probability & Its Applications, 9(1):141–142, 1964.

Public Health Scotland. AE2 - Accident and emergency records, 2020a. https://www.ndc.scot.nhs.uk/National-Datasets/data.asp?SubID=3, Accessed: 6-3-2020.

Public Health Scotland. PIS - Prescribing information systems, 2020b. https://www.ndc.scot.nhs.uk/National-Datasets/data.asp?SubID=9, Accessed: 6-3-2020.

Public Health Scotland. System Watch: urgent care usage, 2020c. https://publichealthscotland.scot/services/ system-watch/#section-1-1, Accessed: 6-3-2023.

Public Health Scotland. SMR datasets - ISD Scotland Data Dictionary, 2023. https://www.ndc.scot.nhs.uk/Data-Dictionary/SMR-Datasets/, Accessed: 6-3-2023.

Geoffrey S Watson. Smooth regression analysis. Sankhyā: The Indian Journal of Statistics, Series A, pages 359–372, 1964.

